# Polygenic Risk Scores and HLA Class II Variants are Biomarkers of Corticosteroid Response in Childhood Nephrotic Syndrome

**DOI:** 10.1101/2025.08.01.25332825

**Authors:** Tiffany Tu, Alejandro Ochoa, Amika Sood, Ashley Drabik, Megan Chryst-Stangl, Brandon Lane, Guanghong Wu, Frank Donovan, Ursula Harper, Settara Chandrasekharappa, Christopher Esezobor, Adaobi Solarin, David Hooper, Christine Sethna, Sandra Amaral, Mahmoud Kallash, Michelle Rheault, Priya Verghese, Vikas Dharnidharka, Eloise Salmon, Patricia Weng, Tarak Srivastava, Michael E. Seifert, Cozumel Pruette, David Selewski, Keisha Gibson, Tracy Hunley, Asiri Abeyagunawardena, Shenal Thalgahagoda, Arvind Bagga, Aditi Sinha, Nicholas Webb, Larry Greenbaum, Ali Gharavi, Krzysztof Kiryluk, Mathias Kretzler, Lisa Guay-Woodford, Simone Sanna-Cherchi, Agnieszka Bierzynska, Ania Koziell, Gavin Welsh, Moin Saleem, Charles Rotimi, Eileen Chambers, Cliburn Chan, CureGN Consortium, PNRC Glomerular disease group, CIBMTR/NMDP Consortium, Annette Jackson, Adebowale Adeyemo, Rasheed Gbadegesin

## Abstract

**Introduction:** Nephrotic syndrome (NS), a common glomerular disease in children, is classified based on response to corticosteroid therapy as either steroid-sensitive nephrotic syndrome (SSNS), or steroid-resistant nephrotic syndrome (SRNS). However, there are currently no reliable predictors of therapy response at initial clinical presentation.

**Methods:** We conducted genome-wide association studies, developed polygenic risk scores (PRS) for therapy response and analyzed classical HLA alleles in 1,997 (994 discovery and 1,003 replication/validation cohorts) previously unstudied children with NS and 3,558 ancestry-matched controls.

**Results:** A significant association with HLA loci defined by variants in *HLA-DQB1, HLA-DRB1*, and *HLA-DQA1* were found for SSNS (but not SRNS), along with a second immune-related SSNS locus: *CLEC16A*. A PRS that discriminates between SSNS and SRNS was validated in two independent cohorts. The HLA haplotype *HLA-DRB1*07:01∼DQA1*02:01∼DQB1*02:02* was associated with ∼4 times the risk of developing SSNS. A model incorporating HLA haplotype, PRS score, and age at onset of the disease was the best predictor of steroid responsiveness with an AUC of 0.68-0.70 and an overall classification accuracy of SSNS versus SRNS of 67-71%.

**Conclusions:** Our findings confirm that SSNS (unlike SRNS) is an immune-mediated HLA-associated disorder. The PRS for therapy response and HLA haplotype can serve as biomarkers and provide a foundation for more accurate diagnoses and tailored and individualized treatment.

**Translational statement:** To identify biomarkers of pattern of steroid responsiveness in childhood-onset nephrotic syndrome, we carried out a case-control study that included over 4,000 samples, including 994 patients with NS in the discovery phase and an additional 1,003 cases from two independent replication cohorts. We identified significant risk of steroid-sensitive NS (SSNS) at HLA class II genes and *CLEC16A* (a gene important in the regulation of T and B lymphocytes). The HLA haplotype *DRB1*07:01∼DQA1*02:01∼DQB1*02:02* was associated with four-fold increased odds of SSNS. A model incorporating HLA haplotype, polygenic risk score, and age at onset of the disease was the best predictor of steroid responsiveness providing useful delineation of steroid sensitivity from steroid resistance. In conclusion, age at onset of disease, HLA class II variants and polygenic risk scores are useful biomarkers of corticosteroid response in childhood NS and may serve as useful clinical decision support tools to guide treatment.

Graphical Abstract

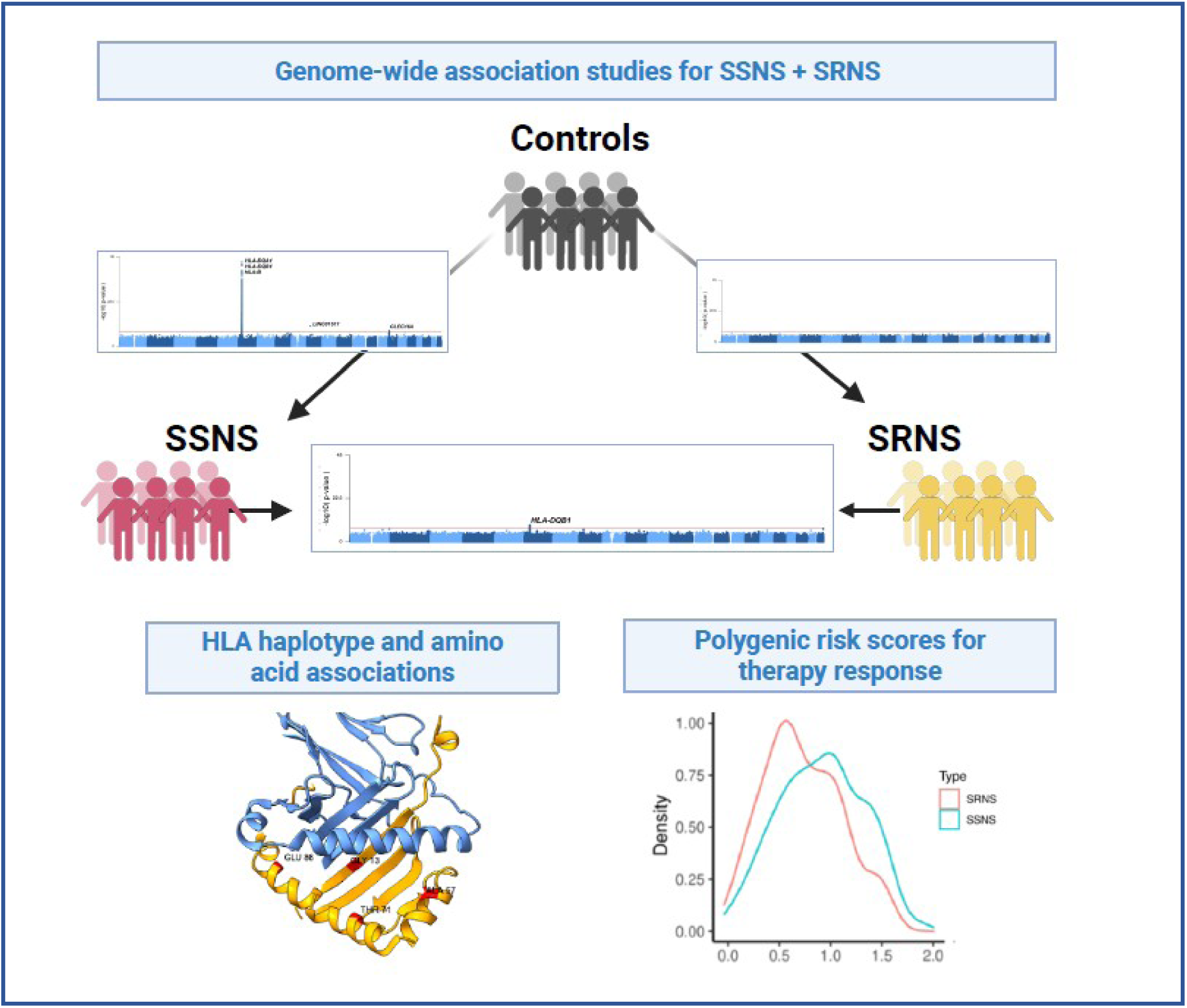

## Introduction

Nephrotic syndrome (NS) is one of the most common pediatric kidney diseases^1^, due to a primary or secondary defect in the glomerular filtration barrier and clinically characterized by massive proteinuria, hypoalbuminemia, and edema. Kidney biopsy morphologic findings in most children with NS are either minimal change disease (MCD) or focal segmental glomerulosclerosis (FSGS)^1^. Approximately 80% of pediatric NS patients respond to steroids, referred to as steroid sensitive NS (SSNS), while 20% are steroid resistant (SRNS)^1^. SRNS progresses more rapidly, often resulting in end stage kidney disease within five to ten years of diagnosis^1^. More than 80 monogenic causes of SRNS have been identified, including *APOL1,* which is associated with FSGS in individuals of African ancestry^2–3^. Nevertheless, the molecular mechanisms of NS remain poorly understood, limiting the development of new diagnostic, prognostic, and therapeutic tools.

Our current knowledge of NS pathogenesis, derived from multiple epidemiologic, clinical, and experimental studies, indicates a critical role for immune dysregulation in T and B lymphocytes in most SSNS and some SRNS patients^4^. Poor response to corticosteroids and other immunomodulating agents in NS patients usually escalates to immunosuppressive agents with major side effects including infections, growth failure, diabetes mellitus, and ocular complications. Thus, exposure to toxic side effects of ineffective therapy is a real risk among NS patients. Currently, there are few reliable risk stratification criteria for disease progression or for determining the immunosuppressive regimen intensity in patients presenting with NS for the first time. Thus, there is a need for robust predictors of therapy response in children with NS to guide therapeutic decisions and minimize unnecessary exposure to futile therapy with serious side effects.

Recent exome- and genome-wide association studies (GWAS) have identified HLA class II variants as primary genetic risk factors for SSNS^5–8^. Although these findings represented important advances, their approaches were limited by at least one of: 1) exclusion of SRNS patients or lack of delineation between SSNS and SRNS, precluding the identification of therapy response-associated variants, 2) use of single-ancestry cohorts that limit generalization to other populations, and 3) omission of age group in the phenotype definition, which may confound results since NS clinical course differs between children and adults.

In this study, we performed a comprehensive analysis of the genetic architecture of pediatric NS. Using a large multi-ancestry cohort with well-defined SSNS, SRNS, and control participants, we performed GWAS to identify significant loci associated with SSNS and with SRNS, overall and in specific ancestries. We confirmed the chromosome 6 HLA region as a robust locus for SSNS and identified an HLA haplotype strongly associated with SSNS. We discovered a chromosome 16 locus in *CLEC16A,* encoding a protein involved in HLA trafficking, and replicated it in an independent cohort. We also identified new suggestive loci on chromosomes 7, 8, 10, and 15. Using this data, we generated and validated polygenic risk scores (PRS) for predicting therapy response (SSNS versus SRNS). Additionally, we analyzed HLA classical alleles and ancestry-specific associations. A model incorporating HLA haplotype, PRS score, and age at onset of the disease distinguishes steroid responsiveness from steroid resistance with an AUC of 0.67-0.70. Our findings lay a foundation for accurate diagnosis and individualized immunomodulation therapy for children with NS.

## Methods

### Discovery cohort

Discovery has 994 multi-ancestry NS cases enrolled in the Duke Genetics of Kidney Disease (Duke GKD; Pro00014951) and Genomics of Nephrotic Syndrome studies (Pro00106061; **Figure 1)**. Written informed consent and/or assent was obtained from all participants. Institutional Review Board approval was obtained from Duke University Medical Center (Durham, NC, USA) and all collaborating institutions. Pediatric NS was defined as age at onset of disease <=21 years. NS was defined as proteinuria (>2+ protein on dipstick, or spot urine protein (mg)/creatinine (mg) ratio >2), hypoalbuminemia (serum albumin <3.5 mg/dl) and edema. Participants with secondary NS were excluded. Clinical data obtained included demographics (age, sex, race/ethnicity), family history of kidney disease, age at onset of symptoms, and laboratory data including urinalysis, urine protein/creatinine ratio and serum creatinine **(Supplementary Table S1)**. Treatment received and pattern of therapy response were classified following KDIGO guidelines^9^.

**Figure 1:**
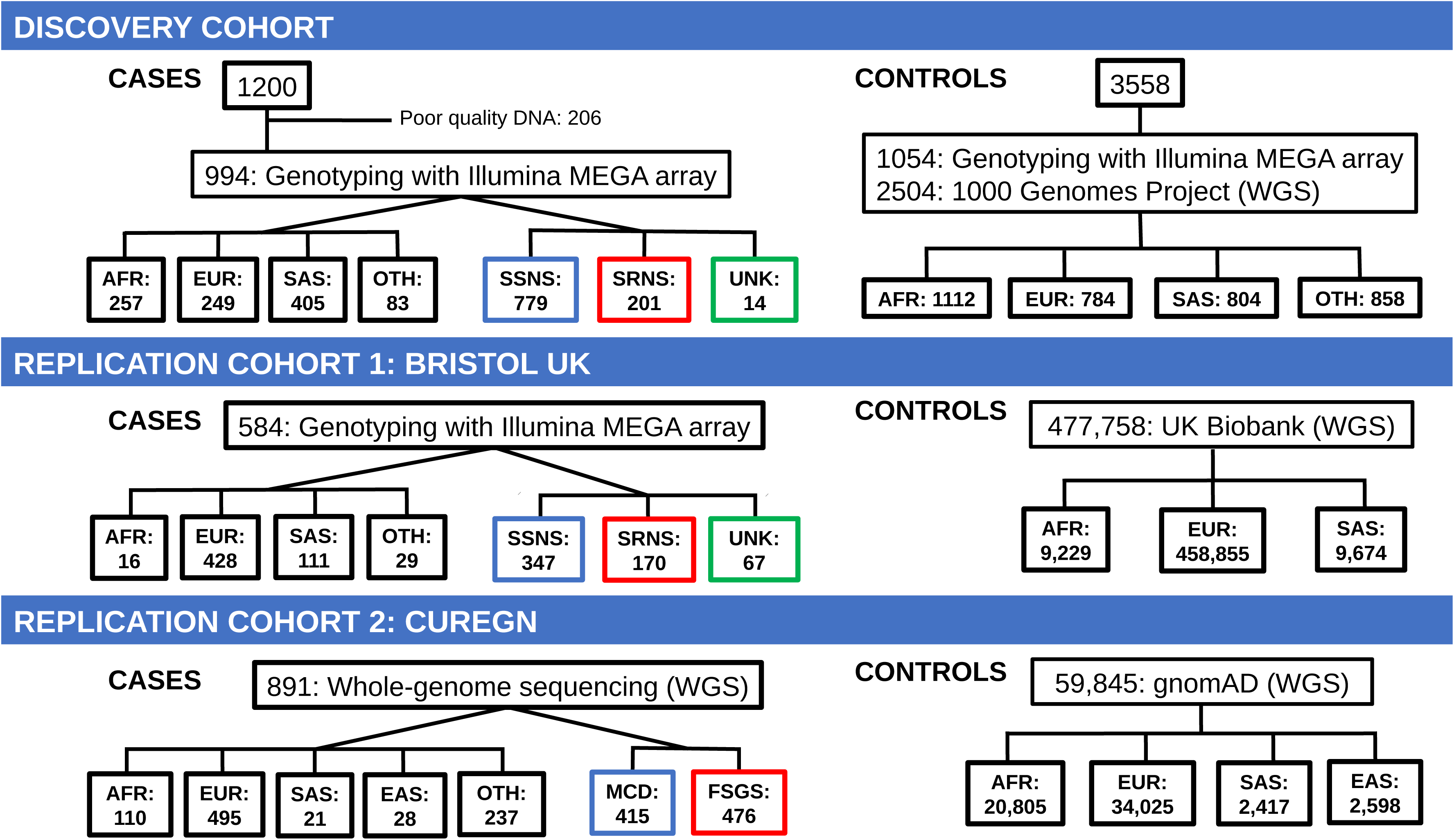
Study design and flow chart. Flow chart for discovery and replication studies showing ancestry of all cases and controls and therapy response by cohort. SSNS: Steroid sensitive nephrotic syndrome, SRNS: Steroid resistant nephrotic syndrome, UNK: Unknown, AFR: African ancestry, EUR: European ancestry (excluding Finnish ancestry for external data), SAS: South Asian ancestry, EAS: East Asian ancestry, OTH: Other ancestries, MCD: Minimal change disease, FSGS: Focal and segmental glomerulosclerosis, WGS: whole-genome sequencing, gnomAD: Genome Aggregation Database.

Discovery controls include: 1) 1,054 adults from the Center for International Blood and Marrow Transplant Research (CIBMTR)/National Marrow Donor Program (NMDP) Research Sample Repository and Duke GKD study controls. Using adults without renal disease minimizes misclassification, since age-matched controls may include cases yet to develop disease^5^. 2) All 2,504 individuals from the 1000 Genomes Project (1000GP)^10^, justified because NS is ultra-rare in the general population (estimated prevalence 2-7/100,000), unlikely to be enriched in a population reference panel not ascertained for kidney disease like 1000GP^1^. The discovery GWAS of 994 NS cases and 3558 controls has 80% power to detect a genotypic risk ratio of 2.0 with minor allele frequency (MAF) of 0.05, or a genotypic risk ratio of 1.7 at MAF=0.1 (**Supplementary Figure S1**).

SNP genotyping (n=2,048) was performed at the National Human Genome Research Institute Genomics Core using the Illumina Infinium® Multi-Ethnic Global Array. Quality control filters resulted in 833,047 SNPs and 1,981 individuals, including 932 cases (725 SSNS, 193 SRNS, 14 unclassified NS) and 1049 controls (**Supplementary Methods, Supplementary Figure S2**). Platform-specific bias filters were applied before merging 1000GP and array data (**Supplementary Methods, Supplementary Figures S3-S4)**, which were then imputed using the TOPMed imputation server^11–12^, retaining only biallelic loci with minor allele count (MAC) ≥20 and imputation R^2^≥0.3.

### Independent Replication Studies

*Bristol, UK*: 584 multi-ancestry pediatric NS patients enrolled at the University of Bristol (under Pro00106061) were genotyped on the same array and lab, and imputed, as the discovery cohort **(Supplementary Figure S5)**. As controls, we used ancestry-specific allele counts from the UK Biobank after QC^13^ (**Supplementary Methods**).

*CureGN:* 891 multi-ancestry NS patients (419 pediatric) enrolled in the CureGN study^14^ **(Supplementary Figure S6, Supplementary Methods).** Whole-genome sequencing data was filtered to retain biallelic SNPs with filter=“PASS”, MAC≥20, and missingness <0.1. As controls, we used ancestry-specific allele counts from gnomAD^15^ after QC (**Supplementary Methods**).

### Statistical analysis

GWAS of NS (versus controls), SSNS (versus controls), SRNS (versus controls), and SSNS versus SRNS were conducted using the SAIGE logistic mixed model^16^. The additive genetic model included as covariates sex, self-reported race/ethnicity, and the first 10 principal components (PCs) of the genotype matrix. We used conditional analysis of significant loci (p<5×10^-8^) to identify independently significant SNPs. Replication tests were done for SNPs with discovery suggestive p<10^-5^, using SAIGE with covariates for individual-level data, or with binomial allele frequency tests for control allele counts. The replication p-value threshold of 0.05 was Bonferroni-adjusted for the number of independent tests determined by linkage disequilibrium (LD) clumping (**Supplementary Methods**).

PRS were developed using *LDpred2* (R package bigsnpr)^17–18^ using three independent genotype datasets named “base”, “training”, and “testing”. Base is used to generate GWAS summary statistics (coefficients) and LD matrices. Training is used to fit PRS parameters that modify coefficients (*LDpred2* grid parameters are heritability, causal variant proportion, and sparseness), to maximize squared partial correlation (R^2^) between PRS and trait conditioned on the top 10 PCs. Training performance is overstated due to overfitting, so final R^2^ and odds ratios (OR) are calculated on the testing data **(Supplementary Methods)**. SSNS-vs-Control were superior base data than SSNS-vs-SRNS for predicting SSNS-vs-SRNS in testing **(Supplementary Figure S7)**. Afterwards, we split discovery into a training set with all 193 SRNS samples and random ancestry-matched 193 SSNS patients, and a base dataset with all 3553 controls and all remaining SSNS patients (532); Bristol and CureGN were used for testing, separately and combined.

The *APOL1* high-risk haplotypes G1 and G2 were determined by rs73885319 (chr22:36265860:A:G) allele G and rs71785313 (chr22:36265995:AATAATT:A) deletion allele, respectively. We associated high risk genotypes (G1G1, G2G2, G1G2) to SSNS or SRNS versus controls using SAIGE.

Imputation of classical HLA alleles (at four-digit/two-field resolution) and amino acid residues was done using the Michigan Imputation Server and the Multi-Ancestry HLA Reference Panel v2.0, which gives high accuracy (94-98%) at G-group resolution in multiple populations^19–20^ (**Supplementary methods)**.

We explored using HLA risk haplotype, PRS, and age as screening tools to distinguish SRNS from SSNS. The sensitivity (probability that test is positive in an individual with SSNS), specificity (probability that test is negative in an individual without SSNS), positive predictive value (probability that an individual with a positive test has SSNS) and negative predictive value (probability that an individual with a negative test does not have SSNS) were computed for each model after logistic regression. Overall performance was evaluated using the area under the receiver operator characteristic curve (AUC) and overall accuracy.

## Results

### Associations for nephrotic syndrome and therapy response

The discovery cases comprised 994 pediatric NS participants, including 257 (25.9%) African, 249 (25.1%) European, 405 (40.7%) South Asian and 83 (8.3%) mixed/other ancestry individuals **(Figure 1)**. The male: female ratio was 1.7: 1, consistent with previous publications^1^. Most patients had SSNS (78.4%), 20.2% had SRNS; response was unknown in 1.4% of participants **(Supplementary Table S1)**. The median age at diagnosis was 3 years (range 1-19 years). Children with SSNS were younger than those with SRNS (median [range] 3.5 [1-19] years versus 7.0 [1-18] years). The 1,054 directly genotyped discovery controls primarily represented African, European, and South Asian ancestries. Using 1000GP as additional controls, we achieved 3,558 controls and a case: control ratio of ∼1: 4 in the overall sample and at least 1:2 in the major ancestry groups. Cases and controls clustered closely after platform harmonization and imputation **(Supplementary Figure S8)**.

The NS discovery GWAS displays a prominent chromosome 6 HLA locus and several secondary loci **(Figure 2A, Supplementary Table S2)**; this association profile is driven primarily by SSNS **(Figure 2B)**. The HLA locus SSNS association replicates previous results^5–8^ **(Figure 2B**, **Table 1)**. The leading HLA SNP is rs17843604, an intronic variant in *HLA-DQA1* (OR=2.57, 95%CI 2.25-2.94, p=2.8×10^-43^; **Figure 2C**). Conditioning on the top SNP identifies rs3129713 upstream of *HLA-DQB1* as a second independent variant at the locus. A second conditional analysis identified a third independent locus, rs113688927 upstream of *HLA-B*. Significant loci outside of chromosome 6 were found on chromosome 16 (lead SNP rs12925642, intronic variant in *CLEC16A*; **Figure 2D**) and chromosome 10 (lead SNP rs114032596, intronic variant in *LINC01517*). Significant SNPs for SSNS are shown in **Table 1** and suggestive SNPs are annotated in **Supplementary Data S1** and summarized in **Supplementary Table S3.**

**Figure 2:**
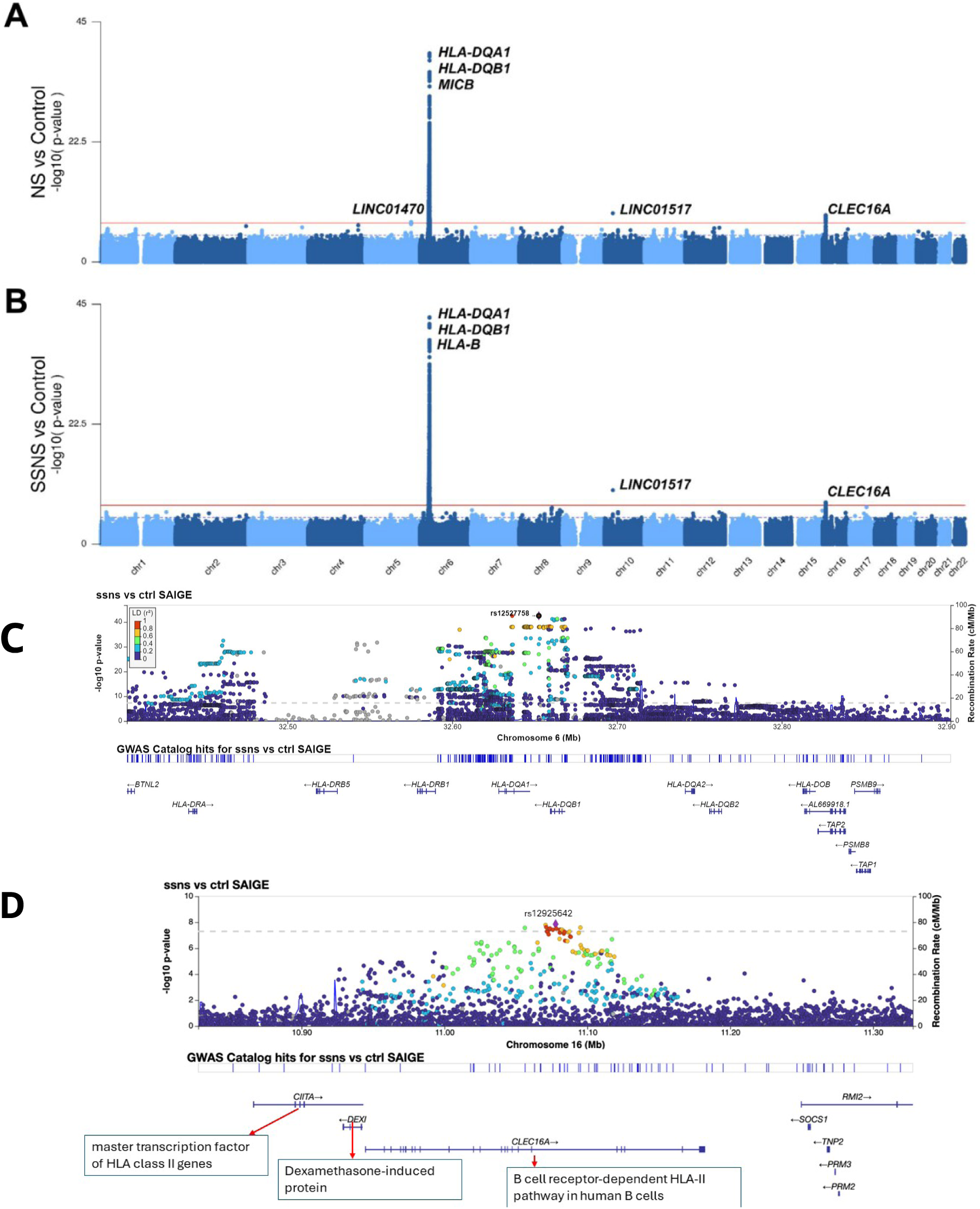
Genome-Wide Association Study (GWAS) Manhattan plots for **(A)** All Nephrotic syndrome (NS) versus controls, showing genome wide significant loci on chromosomes 5, 6, 10 and 16. The chromosome 6 HLA region is the most robust locus with allelic p=7.7×10^-40.^ **(B)** Steroid sensitive nephrotic syndrome (SSNS) versus controls showing significant loci on chromosomes 6, 10, and 16, with allelic p=2.8×10^-43^ for the chromosome 6 region. Regional plots for **(C)** HLA locus for SSNS and **(D)** Chromosome 16 *CLEC16A* locus for SSNS.

**Table 1:**
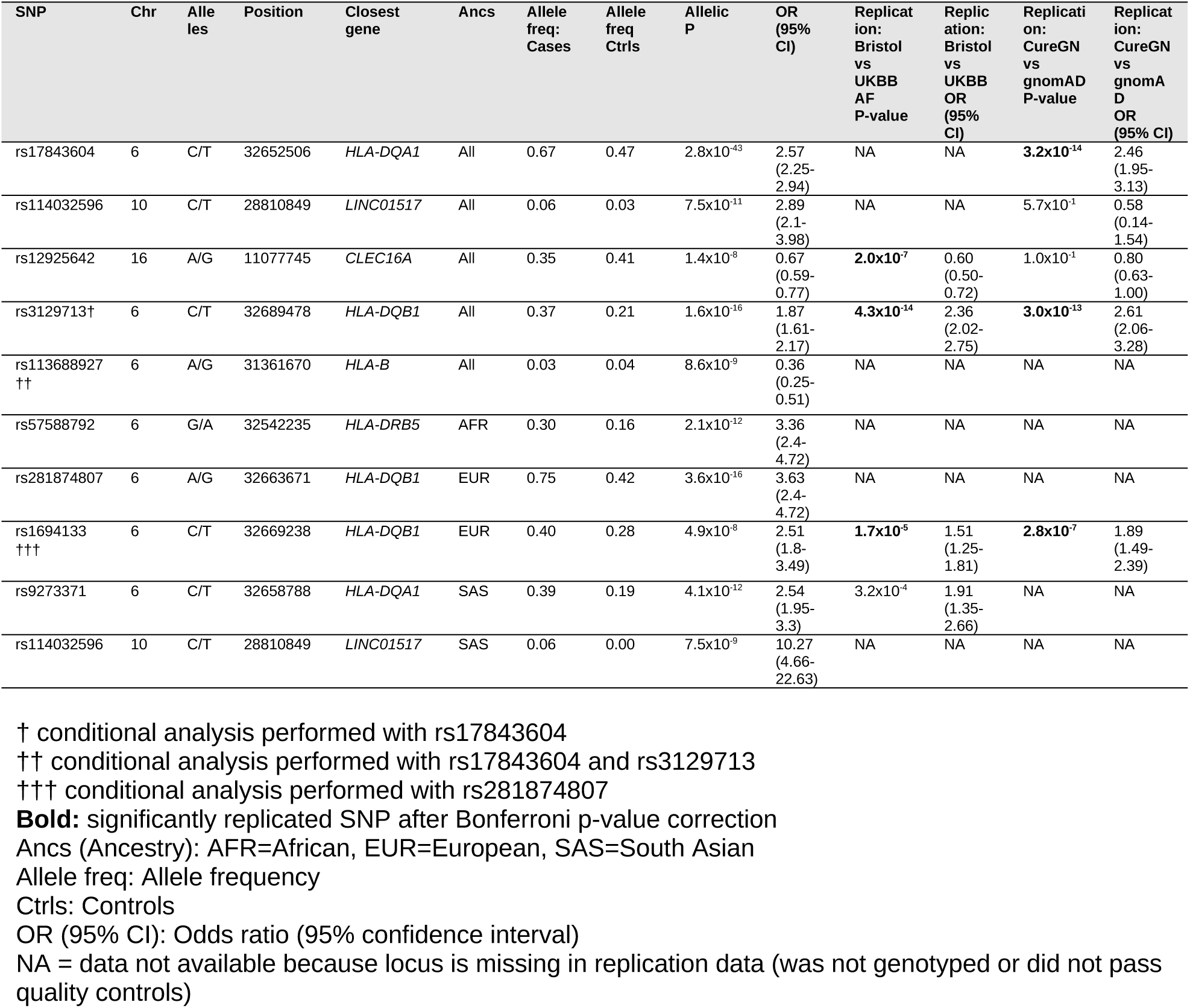
Genome wide significant variants associated with SSNS.

To identify potential population-specific SSNS loci, we performed stratified analysis on the major ancestry groups (African, European, and South Asian): all share significant HLA associations (**Supplementary Figure S9)**. Leading SNPs are rs57588792 (intronic variant in *HLA-DRB5*), rs281874807 (upstream of *HLA-DRB1*) and rs9273371 (nearest gene *HLA-DQA1*) in the African, European, and South Asian cohorts, respectively. Conditional analysis identifies rs1694133 (upstream of *HLA-DQB1*) as independently significant in Europeans. We also identified rs114032596 on chromosome 10 (intronic variant in *LINC01517*) as a risk locus in South Asians. Trans-ancestry meta-analysis did not identify additional loci (**Supplementary Figure S10**). Among significant loci, we replicated rs12925642 (chromosome 16), rs3129713 (chromosome 6), and rs1694133 (chromosome 6) in the Bristol cohort and rs17843604 (chromosome 6), rs3129713 (chromosome 6), and rs1694133 (chromosome 6) in CureGN (**Table 1**).

The SRNS GWAS did not identify significant loci (**Supplementary Figure S11**), but suggestive loci (p<5×10^-6^) appear on chromosome 22, all in *APOL1*. The suggestive *APOL1* variant rs58384577 was replicated in CureGN (**Supplementary Table S4**).

Lastly, we tested for association with steroid responsiveness in NS using SRNS as cases (n=193) and SSNS as controls (n=725). The HLA region had significant variants (*HLA-DQB1* variant rs9274639, OR=0.38, 95%CI 0.27-0.52, p=2.1×10^-9^; **Supplementary Figure S11**), which replicated in Bristol (OR=0.40, 95%CI 0.27-0.59, p=2.72×10^-6^) and CureGN (OR=0.33, 95%CI 0.23-0.49, p=2.95×10^-8^). Overall, associations differ between SSNS and SRNS, driven primarily by HLA class II loci.

### PRS prediction of steroid responsiveness

To predict steroid responsiveness in NS, we developed a PRS for SSNS versus SRNS. The SSNS-versus-SRNS GWAS has a smaller sample size (limited power), so we also considered the larger SSNS-versus-controls GWAS, which more confidently identifies the HLA variants associated with SSNS but not SRNS. Our evaluation determined that scores derived from SSNS-versus-control better predict therapy response (R^2^=0.053, 95%CI 0.023-0.102 for grid method) than those of SSNS-versus-SRNS (R^2^=0.036, 95%CI 0.01-0.073 for grid method), although both perform remarkably well **(Supplementary Figure S7A)**. Henceforth, for PRS construction from SSNS-versus-controls GWAS we reorganized our base, training, and testing datasets to use all samples: we split discovery SSNS samples so most are used with all discovery controls as base data, while the rest of SSNS samples are used with all discovery SRNS samples as training data, matched 1:1 by ancestry; the replication Bristol and CureGN cohorts are used as testing datasets. PRS training results were consistent across all base and training setups **(Supplementary Methods, Supplementary Figures S12-S15)**. The optimal LDPred2 grid method has in Bristol R^2^=0.053, 95%CI 0.023-0.096, in CureGN R^2^=0.068, 95%CI 0.029-0.123, and combined (by meta-analysis) R^2^=0.063, 95%CI 0.036-0.096 **(Figure 3, Supplementary Figure S7B)**. This is noteworthy because we treated CureGN pediatric MCD and FSGS as clinicopathologic correlates for SSNS and SRNS, respectively, so our PRS also distinguishes between these histopathologic NS variants. The best PRS model’s (LDPred2 grid; **Supplementary Data S2**) upper quartile discriminates between SSNS and SRNS patients relative to the lower quartile: in the combined Bristol and CureGN, OR=3.57, 95%CI 2.40-5.38, p=1.62×10^-10^ **(Figure 3)**. The popular “clumping and thresholding” (CT; **Supplementary Data S3**) model has only eight loci, all in the HLA region, and also performs very well **(Supplementary Figure S16)**.

**Figure 3:**
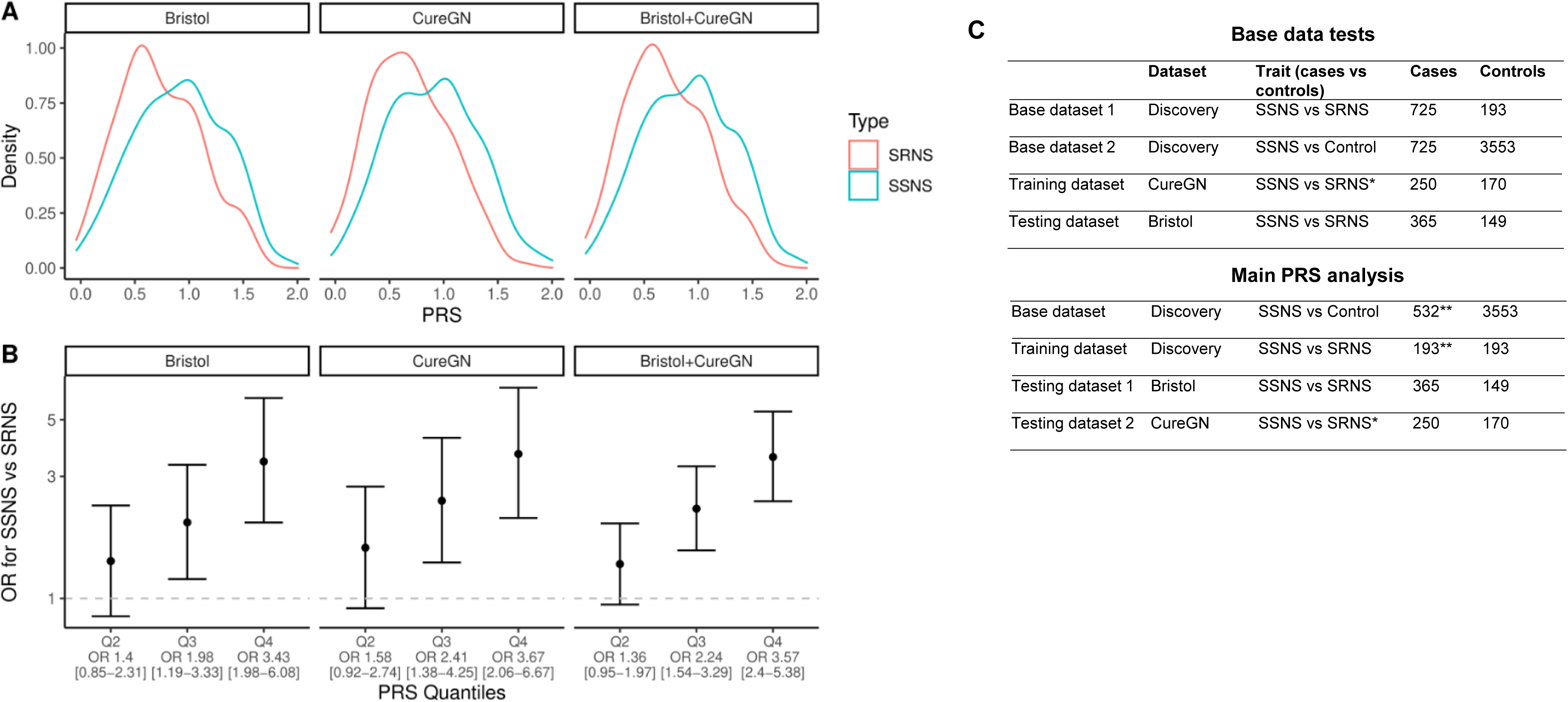
Polygenic risk scores (PRS) predict treatment response in nephrotic syndrome. The PRS was constructed using the LDPred2 grid model from SSNS-versus-Control base data and trained on SSNS-versus-SRNS data, both non-overlapping subsets of the discovery dataset. The test datasets Bristol and CureGN cohorts are also disjoint from each other and the training data. In Bristol cohort, the true diagnoses of SSNS and SRNS are used, whereas for CureGN pediatric MCD and FSGS were treated as pathologic correlates of SSNS and SRNS, respectively; the PRS discriminates these cases in both datasets with similar profiles. **(A)** Kernel density plots of the PRS for the true classes show the overlap and mean shifts, with higher scores for SSNS patients, which are consistent in both test datasets. **(B)** Odds ratios (OR) and 95% confidence intervals (CIs) for SSNS versus SRNS calculated per PRS quartile (ranges shown for reference), which also demonstrate consistent effect sizes across test datasets. The combined “Bristol + CureGN” case has narrower 95% CIs due to its larger sample size. **(C)** Polygenic Risk Score base, training, and testing datasets. **In main PRS analysis, the 725 original Discovery SSNS cases were split into two non-overlapping sets, one for the base GWAS data and the other to train PRS models.

### *APOL1* high risk variants association in African ancestry

We tested the association between SSNS and SRNS and *APOL1* high risk genotypes for kidney disease (G1/G1, G2/G2, G1/G2) in African ancestry. We found a significant association for SRNS (frequency 36.6% in SRNS versus 16.8% in controls; OR=2.62, 95%CI 1.31-5.25; **Supplementary Table S5**), but not for SSNS (frequency 23.6% in SSNS versus 16.8% in controls; OR=1.04, 95%CI 0.66-1.65). This confirms our previous observations that *APOL1* is a risk locus for SRNS but not SSNS^21^.

### HLA haplotype and amino acid associations in SSNS

The most significant 4-digit HLA associations by ancestry included HLA-DRB1*07:01, HLA-DQA1*02:01, and HLA-DQB1*02:02, **(Figure 4, Supplementary Table S6)**. Given strong LD within the HLA region, we explored the hypothesis that HLA haplotypes explain associations better than individual alleles. We tested haplotypes based on HLA-DRB1, HLA-DQA1, and HLA-DQB1 for association with SSNS. We identified both shared and ancestry-specific haplotype associations **(Table 2, Supplementary Table S7**). Haplotypes DRB1*07:01∼DQA1*02:01∼DQB1*02:02 and DRB1*03:01∼DQA1*05:01∼DQB1*02:01 were associated with SSNS in all three ancestry groups. Across the entire sample, the most significantly associated risk haplotype was DRB1*07:01∼DQA1*02:01∼DQB1*02:02, which appears in 25% of cases versus 8% of controls and confers almost 4 times the risk of developing SSNS (OR=3.8, 95%CI 3.3-4.4). Another associated risk haplotype was DRB1*03:01∼DQA1*02:01∼DQB1*02:01 with 11% frequency in SSNS. The most significant protective haplotype was DRB1*15:01∼DQA1*01:02∼DQB1*06:02, which appears in 1% of cases versus 5% of controls (OR=0.17, 95%CI 0.1-0.3, p=2.6×10^-12^). To validate our HLA allele imputations, we performed deep sequencing for 93 participants and compared HLA-DRB1, HLA-DQA1, and HLA-DQB1 alleles and common haplotype associations and obtained 100% concordance.

**Figure 4:**
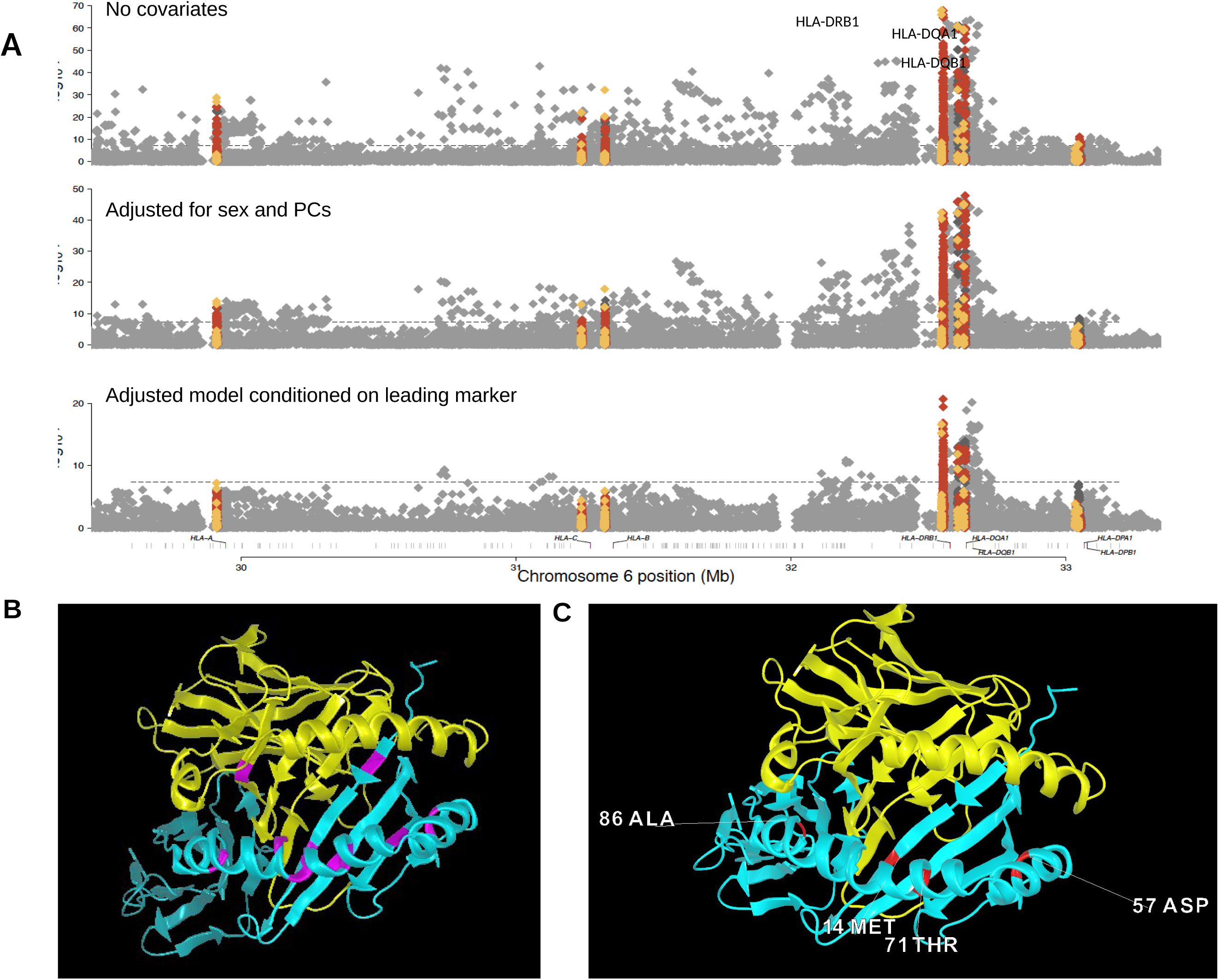
**(A)** HLA regional association plot for SNPs within classical HLA loci **(B)** The HLA-DQA/DQB peptide binding groove showing polymorphic positions (pink) that differ between non-risk allele DQA1*01:02∼DQB1*06:02 and risk allele DQA1*02:01∼DQB1*02:02 **(C)** The most significant amino acid positions (in red) in each ancestry group [beta chain position 71 (South Asian), position 57 & 86 (European), and position 14 (African)], are all in the peptide binding groove and pockets.

**Table 2:** Association of classic 3D HLA haplotype with SSNS.

| **DRB1~DQA1~DQB1** | **OR** | **OR**<br>**Lower**<br>**limit CI** | **OR**<br>**Upper**<br>**limit CI** | **P value** | **Freq.**<br>**controls** | **Freq.**<br>**cases** | **\*Significant in**<br>**which**<br>**populations** |
| --- | --- | --- | --- | --- | --- | --- | --- |
| **07:01~02:01~02:02** | 3.80 | 3.27 | 4.41 | $1.6 \times 10^{-79}$ | 0.08 | 0.25 | All |
| 15:01~01:02~06:02 | 0.17 | 0.09 | 0.3 | $2.6 \times 10^{-12}$ | 0.05 | 0.01 | SAS, EUR |
| **04:01~03:01~03:02** | 2.61 | 1.81 | 3.73 | $1.9 \times 10^{-8}$ | 0.01 | 0.03 | SAS, EUR |
| **03:01~05:01~02:01** | 1.68 | 1.39 | 2.03 | $3.0 \times 10^{-8}$ | 0.07 | 0.11 | All |
| 13:01~01:03~06:03 | 0.25 | 0.14 | 0.44 | $9.0 \times 10^{-8}$ | 0.04 | 0.01 | SAS, EUR |
| 15:03~01:02~06:02 | 0.42 | 0.27 | 0.64 | $2.4 \times 10^{-5}$ | 0.04 | 0.02 | AFR |
| 12:02~06:01~03:01 | 0.29 | 0.14 | 0.55 | $5.7 \times 10^{-5}$ | 0.02 | 0.01 | SAS |
| 09:01~03:02~03:03 | 0.38 | 0.22 | 0.62 | $6.9 \times 10^{-5}$ | 0.03 | 0.01 | SAS |
| 01:01~01:01~05:01 | 0.52 | 0.36 | 0.75 | $2.3 \times 10^{-5}$ | 0.05 | 0.02 | SAS |
| 13:02~01:02~06:04 | 0.33 | 0.14 | 0.66 | $1.3 \times 10^{-3}$ | 0.02 | 0.01 | SAS |
| **07:01~02:01~03:03** | 1.54 | 1.17 | 2.02 | $1.3 \times 10^{-3}$ | 0.03 | 0.05 | None |

Note: Haplotypes with frequencies > 1% shown. Most common risk haplotypes for SSNS shown in **bold**, the other haplotypes are protective of SSNS.
AFR=African, EUR=European, SAS=South Asian

We next constructed extended haplotypes that include classical HLA alleles in A∼B∼C∼DRB1∼DQA1∼DQB1 and tested them for association. Ten extended haplotypes are associated with SSNS, but five of these included the most significant 3-locus haplotype DRB1*07:01∼DQA1*02:01∼DQB1*02:02 **(Supplementary Table S8)**. Therefore, the 3-locus haplotype gives the most parsimonious explanation for HLA haplotype risk.

The most significant amino acid positions associated with SSNS by ancestry are shown in **Supplementary Table S9**. After conditional analysis and Bonferroni correction, we found three independent significant amino acid positions in HLA-DQB1 and HLA-DRB1 in South Asian ancestry (AA_DQB1_71, AA_DRB1_32 and AA_DRB1_233), two in European ancestry (AA_DQB1_57 and AA_DRB1_86), and one in African ancestry (AA_DRB1_13; **Figure 4**). Of note, among all markers (SNPs, HLA alleles, amino acids) in the HLA region, the most significantly associated marker was an amino acid position. The most significant amino acid positions in each ancestry group [beta chain pos 71 (SAS), pos 57 & 86 (EUR) and pos 14 (AFR)], are in key positions of the HLA-DQ/DR peptide binding groove **(Supplementary Table S10)**^22^. This analysis helped refine the HLA association to functional HLA alleles, HLA haplotypes and amino acid residues in multiple ancestries.

### Clinical and genetic predictors of steroid responsiveness

We evaluated using the top HLA haplotype and PRS as screening tools for steroid responsiveness, alone or in combination with age and genotype PCs in the model (**Supplementary Table 11**). In the Bristol and CureGN independent test datasets, the HLA haplotype alone had an AUC of 0.57 and 0.62, respectively, while the PRS had higher AUC between 0.62-0.65. The model incorporating the HLA haplotype, PRS-LDPRED, age and PCs gave the best performance (Bristol AUC 0.68, accuracy 71%; CureGN AUC 0.70, accuracy 67%; **Figure 5**).

**Figure 5:**
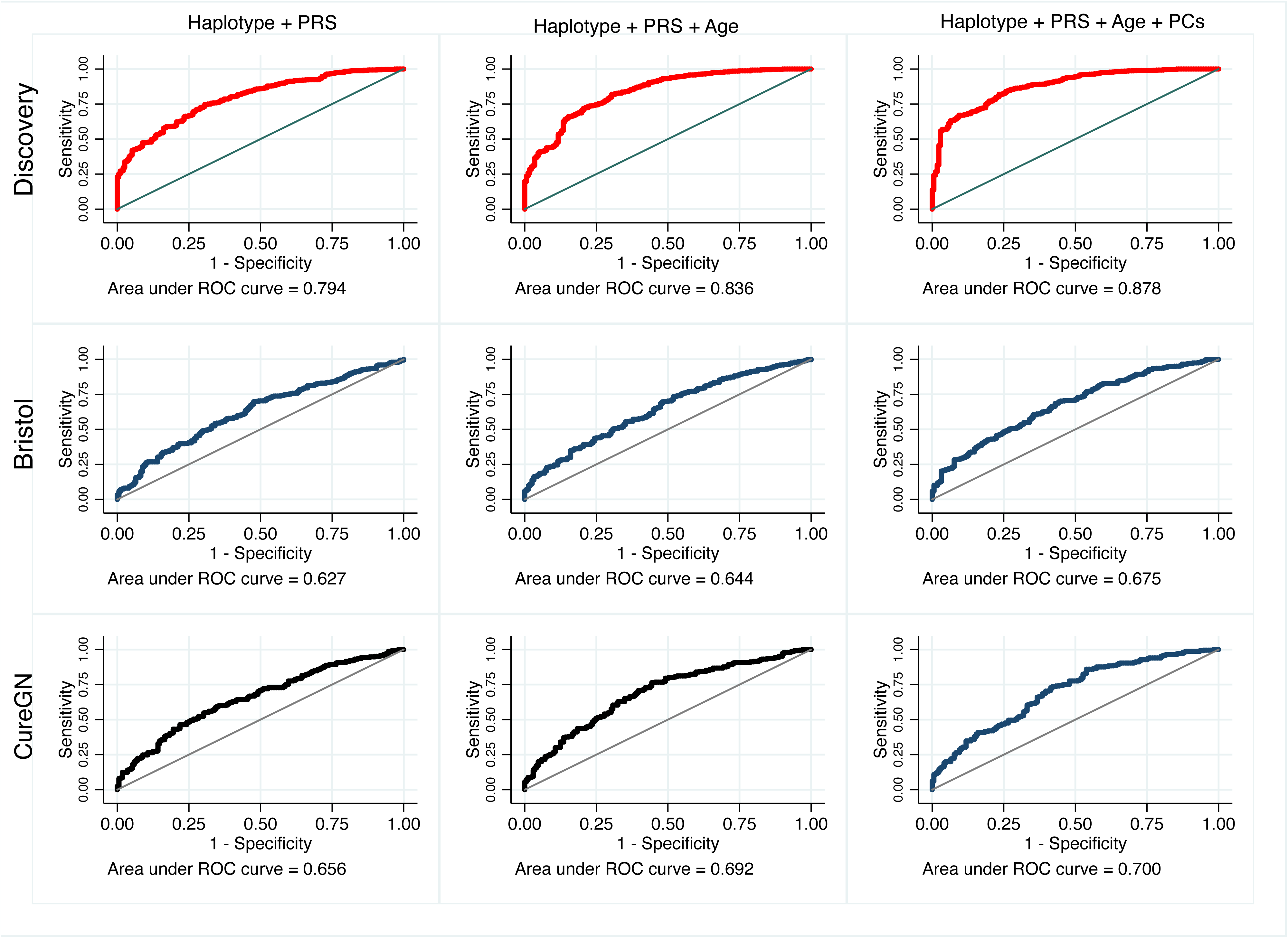
Receiver operator characteristic (ROC) curves of top prediction models of steroid responsiveness. Columns: the top three models, combining the top 3-locus HLA haplotype associated with SSNS, the top PRS model (LDpred2 grid), and optionally age and the top 10 PCs. Rows: models are evaluated on three datasets, discovery (where PRS was trained, so performance is overstated), and the independent Bristol and CureGN datasets (where PRS was not trained, which reflects real world performance of these models).

## Discussion

Herein we report genome-wide analyses to uncover the genetic architecture of NS and predict therapy response. Our cohort is unique because it is a) the largest jointly analyzed (not meta-analyzed) multi-ancestry NS cohort, b) enriched for African and South Asian ancestry compared to previous reports^5–8^, and c) the only one with both SSNS and SRNS patients^5–8^. We replicated previously reported HLA class II associations, which have the largest effect size and are present across ancestries^5–8^. We refined the HLA locus using haplotypes and amino acid positions, finding a 3-locus HLA II haplotype strongly associated with SSNS across ancestries. We also replicated the *CLEC16A* locus in our discovery and Bristol, UK cohorts^8^. Using the well-documented corticosteroid responses in our cohorts, we generated a PRS for therapy response in children with NS and validated this PRS in two independent cohorts. Thus, PRS and HLA class II variants are biomarkers of steroid responsiveness in childhood NS.

To date, all GWAS have identified HLA class II as risk loci for childhood-onset SSNS, consistent with the role of adaptive immunity in disease pathogenesis. However, the role of adaptive immunity in the more severe SRNS has not been clearly defined, although SRNS is likely more heterogeneous and due to primary or secondary defects in podocytes, or dysregulation of the immune system. Here, we showed that HLA variants significantly distinguish between SSNS and SRNS. HLA genes are highly polymorphic, ancestry specific, and haplotypes instead of individual SNPs may be more predictive of disease association and etiology. Our analysis identified HLA-DRB1*07:01∼DQA1*02:01∼DQB1*02:02 as a pan-ancestry risk haplotype that conferred ∼4 times the risk of SSNS. We also identified a protective HLA-DQA1*01:02∼DQB1*06:02 heterodimer common across ancestries. Furthermore, we identified rare ancestry-specific haplotypes, which may explain differences in SSNS prevalence between ancestries. Further investigation of the effect of haplotype on antigen presentation may yield additional insights to the molecular mechanisms of nephrotic syndrome and identification of novel therapeutic targets.

We identified two significant non-HLA loci of interest. The top one is in chromosome 16, an intronic variant rs12925642 in *CLEC16A* encoding for C-type lectin domain containing 16A, which we replicated in another independent cohort. A recent GWAS meta-analysis first associated a *CLEC16A* variant (rs8062322, in high LD with rs12925642, r^2^=0.70) with pediatric SSNS^8^, making our study its first independent replication. *CLEC16A* variants have also been associated with immune-mediated conditions such as diabetes mellitus, multiple sclerosis and rheumatoid arthritis^23–30^. The variant we found is linked to GWAS variants for eosinophil count, TNFR6 level, and immune mediated diseases such as type 1 diabetes, multiple sclerosis, primary biliary cholangitis, SLE, acne vulgaris, acute interstitial nephritis, IBD among others in the OpenTargets Genetics Database (https://genetics.opentargets.org/Variant/16_11070710_G_C/associations). This variant is also an eQTL for DEXI (Dexamethasone-Induced Transcript or DEXI Homolog) in whole blood, lymphoblastoid cell lines and multiple tissues in GTEx, indicating that it regulates transcript levels of a dexamethasone-induced protein in immune response and other cells. *CLEC16A* is also important in the regulation of B and T cells. One study found that *CLEC16A* participates in the B cell receptor-dependent HLA class II pathway in human B cells, and this regulation is impaired in autoimmune disease such as multiple sclerosis^16^. Another study reported that *CLEC16A* expression is induced in CD4+ T cells upon T cell activation^19^. The low frequency of these variants in the population, their association to SSNS in three independent cohorts, and the role of *CLEC16A* in regulation of immune cells in health and disease together establish *CLEC16A* as a major second SSNS locus acting in concert with HLA loci. The other significant locus in our study is in a long intergenic non-protein coding RNA (LINC01517), with unknown functional significance, and risk variant present in South Asian ancestry only, precluding replication.

We cannot currently predict at initial presentation which NS patients will respond to corticosteroids. All patients are initially treated with corticosteroids, so unresponsive patients are exposed to major toxicity and side effects without therapeutic benefit. To address this problem, we developed a PRS using SSNS and SRNS patients and controls and tested the score in an independent UK cohort of SSNS and SRNS patients. The PRS was able to predict corticosteroid responsiveness in this cohort and the highest quantile PRS was associated with steroid sensitivity (OR=3.57, 95%CI 2.40-5.38). The robustness of the score was confirmed by predicting MCD versus FSGS (clinicopathologic correlates of SSNS and SRNS, respectively) in pediatric patients in CureGN. This is the first study to develop a PRS for corticosteroid response in childhood NS or any other chronic kidney disease (CKD). The performance of our PRS is similar or better than PRS for other kidney diseases such as diabetic kidney disease, IgA nephropathy, and multi-etiology CKD^31–33^. We developed this PRS in our multi-ancestry cohort and therefore expect our findings to generalize to most childhood-onset NS patients. Lastly, we showed that combining this PRS with HLA haplotype and age at onset can correctly classify SSNS and SRNS in 67-71% of cases. These predictors for steroid responsiveness can support clinical decisions *at diagnosis* and result in more rational and personalized use of corticosteroid in the management of NS. Prospective confirmation of our findings in combination with clinical predictors of therapy response will enable clinical application of this tool to identify children with NS that are likely to be steroid resistant, minimizing exposure to steroids and its potential complications in those unlikely to benefit from it, and earlier initiation of alternative therapeutic agents like calcineurin inhibitors.

Our study has many significant novel findings and strengths. This is the first study with both SSNS and SRNS patients, which enabled showing that they have distinct genetic profiles. We defined a robust HLA haplotype for SSNS. Since HLA testing is a widely used clinical tool (to investigate suspected immune-mediated or autoimmune disease, for organ transplantation, etc.), the HLA haplotype we identified could be a point-of-care diagnostic tool. We generated the first PRS for therapy response in NS, which with further refinement could inform whether to treat NS with corticosteroid or start with second line agents. Finally, we showed that *CLEC16A* is a robust second locus for SSNS. Further investigation of this target can potentially lead to better understanding of disease pathogenesis and new therapeutic targets. We mitigated against inflated type 1 error due to case/control imbalance, which often affects studies of rare diseases and low frequency variants, by using a logistic mixed-effects model that accounts for this issue.

A weakness of our study is the inability to study endophenotypes such as frequent relapsing, steroid dependent, and secondary steroid resistant forms of NS because of sample size limitations. This weakness limits the application of our potential diagnostic tools in these subgroups of NS patients.

In conclusion, we performed GWAS, PRS, and HLA analyses in patients with childhood NS, including both SSNS and SRNS subtypes. We found a risk HLA haplotype across multiple ancestries, defined PRS for response to corticosteroid in children with NS and identified a robust genetic second locus for SSNS in *CLEC16A*. The study provides compelling evidence of different genetic risk profiles for SSNS versus SRNS, with SSNS being primarily immune-mediated. Steroid responsiveness as a clinical observation reflects these underlying genetic differences. We find that PRS and HLA haplotypes are biomarkers of therapy response in childhood SSNS and provide a solid foundation for clinical translation and personalized treatment of NS.

## Supporting information

List of consortia members

## Data Availability

Genotype and phenotype data generated for this study will be available on the NIH's Database of Genotypes and Phenotypes (dbGaP) and on ImmPort (for samples that consented to such data sharing; both in process), or through collaboration with the corresponding author upon reasonable request. The CureGN study data is available on dbGaP under accession phs002480.v5.p4. The high-coverage version of the 1000 Genomes Project was downloaded from ftp://ftp.1000genomes.ebi.ac.uk/vol1/ftp/data_collections/1000G_2504_high_coverage/working/20190425_NYGC_GATK/. The code generated during this study is available on GitHub at https://github.com/OchoaLab/nephrotic-syndrome/.

## Acknowledgments

We acknowledge all the participants in the study. We appreciate the technical support provided by the Duke Molecular Physiology Institute genomics core and Duke Center for Genomics & Computational Biology.

## List of Supplementary materials

1. Supplementary text: Supplementary methods, Supplementary results, Supplementary Tables, Supplementary Figures (DOCX file)
2. Supplementary Data S1: all GWAS suggestively significant loci annotated tables (XLSX file)
3. Supplementary Data S2: PRS coefficients for ldpred2-grid method (TXT file)
4. Supplementary Data S3: PRS coefficients for CT method (TXT file)
5. List of consortia members (XLSX file)

