## Supplementary Materials for "Polygenic Risk Scores and HLA Class II Variants are Biomarkers of Corticosteroid Response in Childhood Nephrotic Syndrome"

### Supplementary Methods

#### Discovery array genotype data quality control

The array data (2,048 individuals plus 18 intended duplicates, 1,748,250 SNPs) first had duplicate individuals removed. Duplicate individuals are expected to have a kinship coefficient of 0.5 (same as identical twins), while the next degree of relatedness, parent-child and sibling, both have expected kinship is 0.25. Thus, we identified individual pairs as those with kinship greater than 0.354 (the geometric mean of 0.25 and 0.5) using the KING-robust estimator implemented in plink2 (1,2). This process identified both intended and unintended duplicates. Both individuals in an unintended duplicate were removed if they disagreed in sex, race/ethnicity, or diagnosis; 9 such pairs (18 individuals) were removed. Otherwise, the individual in the pair with the most missingness was removed, resulting in 18 intended plus 34 unintended duplicates additionally removed. Afterwards, the data underwent standard quality control filters: only autosomal biallelic SNPs were kept; 15 individuals with missingness above 10% were removed; variants with missingness above 10%, minor allele frequency below 0.01, or Hardy-Weinberg equilibrium (HWE) test p-value below $1x10^{-4}$, and duplicated loci were removed. This filtering resulted in 1,981 individuals (932 cases and 1049 controls) and 833,047 SNPs. Of the 932 NS cases, we have 725 SSNS, 193 SRNS, and 14 unclassified NS.

#### Identification of ancestry subgroups for harmonization and stratified analysis

An admixture model with K = 5 ancestries--which differentiated African, European, South Asian, East Asian, and Native American ancestries--was fit to the array data using ADMIXTURE (3). For the purpose of stratified analyses, we studied African, European and South Asian ancestry subgroups, corresponding to the 1000 Genomes AFR, EUR and SAS super-populations, respectively. For the allele frequency tests and ancestry sub-analyses only, individuals with less than 80% of the primary ancestry were excluded, resulting in 495 individuals for African ancestry, 486 for European, and 591 for South Asian, respectively (**Supplementary Figure S2**). The ancestry clusters are visualized using principal components analysis (**Supplementary Figure S8A)**.

**Harmonization and merging of discovery array cohort with 1000 Genomes Project**

We use the high-coverage NYGC version of the 1000 Genomes Project (4), which is in the same genome version GRCh38 as our array data. Using plink2 (2) we keep only autosomal biallelic SNP loci with filter "PASS", resulting in 91,784,660 loci and 2,504 individuals.

To address platform-specific issues in merging array genotypes with the 1000 Genomes genotypes, we undertook a series of additional quality control measures. The GWAS array dataset has 762,629 SNPs in common with 1000 Genomes, the remainder were excluded from further analysis. Next, we removed SNPs that are systematically different between array and WGS data, as follows. This test used only control individuals that have more than 80% of each of the three major ancestries: African (301 individuals), European (274), and South Asian (248); in 1000 Genomes, these major ancestries correspond exactly to the super populations AFR (661 individuals), EUR (503), and SAS (489), respectively. The data were further grouped by genotyping platform (array vs WGS). We performed a likelihood ratio test for each SNP, which models allele counts per ancestry and platform as Binomial with some group-specific allele frequency, and tested the null hypothesis that allele frequencies for each ancestry are equal between array and WGS, as expected since both groups are controls (under the alternative hypothesis, allele frequencies are different between platforms for at least one ancestry). Each ancestry is treated as independent data, and the resulting log-likelihood ratio statistic has a Chi-squared distribution with 3 degrees of freedom (number of ancestries), which was used to calculate p-values. We removed significant SNPs with $p < 1x10^{-10}$. Additionally, for SNPs whose reference and alternate alleles are reverse complements of each other, we flip alleles (assumed to be the wrong strand) if the reverse orientation has a greater p-value; those SNPs are removed if the p-values of both orientations are significant. P-value distributions are available in **Supplementary Figure S3**, and allele frequency plots showing SNP classifications are in **Supplementary Figure S4**. In total, 1,263 SNPs were removed and 3,178 were flipped, the remainder were unchanged. The merged and filtered array and 1000 Genomes data has a total of 4,485 individuals (932 NS cases, 3,553 controls), and 761,366 SNPs. The merged data included all previous array individuals regardless of ancestry and admixture.

##### Bristol quality control

Cases in this cohort consisted of 588 individuals with nephrotic syndrome enrolled at the University of Bristol. Four samples did not pass quality controls (high missingness), finally leaving 584 for analysis. Admixture analysis done as described earlier resulted in 428 individuals of primarily European ancestry, 111 South Asian, 16 African, and 29 admixed or other ancestries **(Figure 1, Supplementary Figure S5)**. 347 of the participants had SSNS disease course and 170 SRNS, the disease course is unknown in 67 participants **(Figure 1).** The Bristol cohort was genotyped in the same NHGRI Lab using the same Infinium® Multi-Ethnic Global BeadChip that we used for the discovery GWAS. Quality control and imputation was also done using the same procedures.

##### CureGN quality control

This cohort of 891 patients (419 with age of onset of disease <21 years, treated as pediatric) and diagnosis of minimal change disease (MCD: 415 individuals, 47%; 250 pediatric individuals, 59.7%) or focal and segmental glomerulosclerosis (FSGS: 476 individuals, 53%; 169 pediatric individuals, 40.3%) were enrolled as part of the CureGN study. Pediatric MCD and FSGS were treated as SSNS and SRNS, respectively. The study participants underwent whole genome sequencing (WGS). We kept only biallelic SNP loci with filter “PASS”, minor allele count >= 20, and missingness less than 0.1, and lifted over coordinates from hg19 to hg38, resulting in 8,331,142 loci. To perform the admixture analysis, 1000 Genomes was merged with CureGN; the analysis identified no platform-specific genotyping biases in this case, which makes sense since both platforms are WGS. Our admixture analysis identified 495 (55.6%; pediatric 218, 52.0%) individuals of primarily European ancestry, 110 (12.3%; pediatric 64, 15.3%) African ancestry, 21 (2.4%; pediatric 10, 2.4%) South Asian ancestry, 28 (3.1%; pediatric 9, 2.1%) East Asian ancestry, and 237 (26.6%; pediatric 118, 28.2%) admixed **(Figure 1, Supplementary Figure S6).**

##### UK Biobank allele counts

Ancestry-specific allele counts from WGS data (GRCh38 coordinates) were obtained from the freely-available UK Biobank Allele Frequency Browser at https://afb.ukbiobank.ac.uk/, accessed on February 2024. This data was generated by the WGS consortium under the UK Biobank Resource (project ID 52293).

**GnomAD allele counts**

GnomAD version 4.0 WGS data (GRCh38 coordinates) were used to extract ancestry-specific allele counts, which were downloaded as VCF files from their website at https://gnomad.broadinstitute.org/.

**GWAS replication tests using LD clumping, allele counts and quality control**

Replication tests were carried out using the Bonferroni procedure for multiple hypothesis testing. This enables loci to be significant at a reduced nominal level compared to the genome-wide significance level of 5e-8 since the number of replication loci is much smaller than the number of tests in a complete GWAS. This is often necessary as well since replication cohorts can have smaller sample sizes so there is reduced statistical power to replicate. The number of independent tests in the discovery data was determined using the number of clumps calculated using LD clumping, since many significant loci are in LD with each other, particularly in the HLA region. We performed LD clumping of the SNPs in the discovery dataset that satisfy p<1e-5 only, using plink 1.9 using an LD $R^2$ threshold of 0.3, a window size of 6 Mb, a p-value ceiling (--clump-p1) of 1 (no filter), and a secondary p-value threshold (--clump-p2) of 1 (no filter).

Replication tests involving individual data only (SSNS versus SRNS trait) were carried out using SAIGE with covariates, same as for the discovery analysis, using all loci (not just those we sought to replicate) to estimate the genetic relatedness matrix.

For replication tests involving external control allele counts (UK Biobank or gnomAD), we reused the Binomial allele frequency test described above (harmonization and merging with 1000 Genomes), to test for each replication locus whether the allele frequencies in a disease cohort (Bristol or CureGN) were significantly different from those of the control cohort (UK Biobank or gnomAD), using the allele counts of several ancestries as applicable. For Bristol the most prevalent ancestries are African, European, And South Asian, so we paired them with the UK Biobank ancestries African, Non-Finnish European, and South Asian, respectively. For CureGN the most prevalent ancestries are African, European, South Asian, and East Asian, so gnomAD we used the pre-defined African/African-American, Non-Finnish European, South Asian, and East Asian ancestry groups.

For Bristol, before testing a given locus for association (between internal cases and external controls), as an additional quality control we repeated the platform-specific genotyping bias test between internal and external controls, since internal and external data were usually genotyped with different platforms, which could result in false positive replications. Since Bristol cases were genotyped using the same array as the discovery cohort, we reused the discovery control genotypes (after imputation, same as the Bristol samples) and tested whether allele frequencies differ between our discovery controls and the UK Biobank controls: significant cases were determined to exhibit platform biases and were excluded from testing for association. For CureGN, the platform bias test was neither possible (because its controls, 1000 Genomes, were used in the discovery GWAS, so the results would be circular) nor needed, since both it and gnomAD are WGS datasets, and there was no evidence of platform biases in the form of clear false positive or unexpectedly numerous replications.

**Polygenic risk scores (PRS) Methods**

Prior to training the LDPred2 grid method, we considered the LDPred2 “inf” (infinitesimal) model (9,10), whose sole parameter is the heritability. We performed a grid search for the value of the heritability that maximizes $R^2$ in the training data, considering heritability values between 0.01 and 0.1 in increments of 0.01, and between 0.1 and 0.9 in increments of 0.1. In all cases performance was relatively insensitive to heritability, but very low heritability values were optimal by small margins: $h^2 = 0.01$ for both SSNS-vs-SRNS and SSNS-vs-Control discovery base data and CureGN training, and $h^2 = 0.06$ for the base SSNS-vs-Control trained on SSNS-vs-SRNS discovery data (**Supplementary Figure S12**). Thus, small heritability values centered around $h^2 = 0.1$ were considered when training the more complex LDPred2 grid method.

For the LDPred2 grid method (9,10), scores were calculated for all 168 combinations of the following three parameters: heritability ($h^2$) in {0.03, 0.07, 0.1, 0.14}, the 21 proportions of causal variants (p) between 1e-5 and 1 in equal spacing in log scale, rounded ({1e-n, 1.8e-n, 3.2e-n, 5.6e-n} for n between 1 and 5, and also {1}), and sparse or not sparse (dense). In the training data, performance varied by the evaluation, although the models were less sensitive to the heritability and whether the model is sparse or not, and most sensitive to the choice of proportion of causal variants (**Supplementary Figure S13**). For SSNS-vs-SRNS discovery base data and CureGN training, the best model was sparse, with $h^2 = 0.03$, and p = 5.6e-5. For SSNS-vs-Control discovery base data and CureGN training, the best model was dense, with $h^2 = 0.03$, and p = 1e-3. Lastly, for the base SSNS-vs-Control trained on SSNS-vs-SRNS discovery data, the best model was dense, with $h^2 = 0.1$, and p = 5.6e-4.

For the LDPred2 auto method (11), scores were calculated using an initial $h^2 = 0.1$, an initial vector of proportions of causal variants consisting of 30 values between 1e-4 and 0.2 with equal spacing in log scale, and a shrinkage coefficient of 0.95. Genotype coefficients from the posterior distribution with in-sample $R^2$ greater than 0.95 times the 95th percentile were averaged to produce the final model. (The auto method does not require a separate training dataset, so no training results are available.)

For the LASSOsum2 method (12), also implemented in the R package bigsnpr, scores were calculated for all 120 combinations of the following two parameters: the shrinkage parameter delta in {1e-3, 1e-2, 1e-1, 1}, and the penalization coefficient lambda, 30 values where the minimum is 0.01 times the maximum (the precise values are determined dynamically and are data dependent). In the training data, performance varied by the evaluation, although the models were less sensitive to the shrinkage, and most sensitive to the choice of penalization coefficient (**Supplementary Figure S14**). For SSNS-vs-SRNS discovery base data and CureGN training, the best model has delta = 1 and lambda = 0.166. For SSNS-vs-Control discovery base data and CureGN training, the best model has delta = 1 and lambda = 0.123. Lastly, for the base SSNS-vs-Control trained on SSNS-vs-SRNS discovery data, the best model has delta = 1 and lambda = 0.0885.

For the clump and threshold (C+T) method, as implemented in the R package bigsnpr (13), scores were calculated for all 1400 combinations of the following parameters: LD squared correlation threshold (for clumping) in {0.01, 0.05, 0.1, 0.2, 0.5, 0.8, 0.95}, base window sizes of {50, 100, 200, 500} which are divided by the LD threshold to get actual window sizes, and 50 p-value thresholds with equal spacing in log scale between the minimum p-value and the largest p-value or 0.1, whichever is bigger. In the training data, performance depended strongly on both the LD and p-value thresholds (**Supplementary Figure S15**). For SSNS-vs-SRNS discovery base data and CureGN training, the best model has an LD threshold of 0.8, a window size of 625, and a log10(p) threshold of 6.12. For SSNS-vs-Control discovery base data and CureGN training, the best model has an LD threshold of 0.8, a window size of 62, and a log10(p) threshold of 28.4. Lastly, for the base SSNS-vs-Control trained on SSNS-vs-SRNS discovery data, the best model has an LD threshold of 0.1, a window size of 1000, and a log10(p) threshold of 11.9.

Lastly, we also ran a “stacked” variant of C+T, which finds using penalized logistic regression (elastic net) a linear combination of the best C+T models to improve its predictions (13). We used default parameters as implemented in the R package bigsnpr, which results in a single model as output (there is no explicit training of grid parameters, as in the previous cases, that can be visualized given the limited outputs provided).

The testing results show that all of these models attain similarly high performance in predicting steroid therapy response, with mean $R^2$ estimates between 0.04-0.06 and confidence intervals that exclude not only zero but also 0.02 for all but the worst model (using the combined “Bristol+CureGN” as the definitive test results; **Supplementary Figure S7B**). The best performing methods correspond to very different strategies, including LDPred2 grid, LASSOsum, and C+T, while LDPred2 Inf and Auto were the worst relative performers. Overall, these results suggests that steroid therapy response is predictable with various modeling strategies, and that this trait is probably closer to monogenic rather than polygenic (since C+T performs well).

**HLA imputation**

Imputed HLA alleles and amino acids were filtered for frequency >= 1% and high-quality markers ($r^2$ >= 0.7) before analysis. We analyzed two-field (four-digit) HLA alleles representing specific HLA proteins and not serologic antigen groups. Logistic regression was used to test association between the imputed dosages of the variants and SSNS with sex and genotype-derived principal components as covariates. Conditional analyses were conducted by including the most significant variant in the regression model and testing for residual association in the other variants. Given the well-known differences between populations in the MHC region, we consider as our primary analyses the per-population analysis. HLA analyses were done using PLINK 1.09 and the HLA-TAPAS pipeline (2,5). As a sensitivity test, we also did HLA imputation using the HLA Genotype Imputation with Attribute Bagging (HIBAG) R/Bioconductor package and the IKMB multiethnic reference panel (6,7). We used BIGDAWG v2 to construct HLA haplotypes, estimate haplotype frequencies and do haplotype association analysis using the EM algorithm (8). 3D structures were visualized and manipulated using UCSF ChimeraX.

#### Supplementary Tables

##### Supplementary Table S1: Clinical characteristics of the discovery cohort by therapy response

| **Parameters** | **Cohort size: 994 n (%)** |
|---|---|
| Ancestry | |
| African | 257 (25.9) |
| European | 249 (25.1) |
| South Asian | 405 (40.7) |
| Others | 083 (8.3) |
| Sex M:F | 1.7:1 |
| Age at onset of disease in years Median (Range) | 3.0 (1-19) |
| Therapy response | |
| SSNS | 779 (78.4) |
| SRNS | 201 (20.2) |
| Unknown | 014 (1.4) |

#### Supplementary Table S2: Genome wide significant variants associated with all patients with nephrotic syndrome before and after conditional analysis

| SNP | Chrom | Allele A2 | Position | Type | Gene/ Closest gene | Ancestry | Allele frequency cases A2 | Allele Frequency controls A2 | Allelic P-value | OR (95% CI) | Replication: Bristol vs UKBB P-value | Replication: Bristol vs UKBB OR (95%CI) | Replication: CureGN vs gnomAD | Replication: CureGN vs gnomAD OR (95%CI) |
|---|---|---|---|---|---|---|---|---|---|---|---|---|---|---|
| rs17843604 | 6 | C/T | 32652506 | Protein-coding | *HLA-DQA1* | All | 0.65 | 0.47 | $7.7x10^{-40}$ | 2.22 (1.98-2.5) | NA | | $1.55x10^{-16}$ *** | 1.64 (1.46-1.84) |
| rs114032596 | 10 | C/T | 28810849 | non-coding intronic | *LINC01517* | All | 0.054 | 0.03 | $7.4x10^{-10}$ | 2.5 (1.87-3.35) | NA | | $9.14x10^{-1}$ | 1.06 (0.62-1.68) |
| rs11860603 | 16 | T/C | 11071160 | non-coding intronic | *CLEC16A* | All | 0.37 | 0.42 | $1.73x10^{-9}$ | 0.69 (0.61-0.78) | $1.25x10^{-4}$ | 0.76 (0.67-0.87) | $2.48x10^{-1}$ | 0.89 (0.79-1.00) |
| rs567166387 | 5 | A/T | 152964673 | non-coding intronic | *LINC01470* | All | 0.016 | 0.01 | $3.24x10^{-8}$ | 4.55 (2.66-7.79) | NA | | NA | |
| rs3129713† | 6 | C/T | 32689478 | None | *HLA-DQB1* | All | 0.34 | 0.21 | $5.21x10^{-15}$ | 1.71 (1.5-1.96) | $2.64x10^{-10}$ *** | 1.66 (1.47-1.88) | $2.01x10^{-11}$ *** | 1.64 (1.43-1.87) |
| rs2855812†† | 6 | G/T | 31504943 | 5upstream, intronic | *MICB* | All | 0.30 | 0.21 | $1.90x10^{-8}$ | 1.47 (1.28-1.68) | $1.31x10^{-13}$ *** | 1.67 (1.45-1.91) | $3.52x10^{-1}$ | 1.12 (0.99-1.27) |
| rs57588792 | 6 | G/A | 32542235 | None | *HLA-DRB5* | African | 0.28 | 0.16 | $4.86x10^{-12}$ | 2.92 (2.15-3.96) | NA | | NA | |
| rs9394106 | 6 | A/G | 32652455 | None | *HLA-DQA1* | European | 0.62 | 0.42 | $4.30x10^{-13}$ | 2.43 (1.91-3.09) | NA | | $2.83x10^{-31}$ *** | 2.13 (1.88-2.43) |
| rs9270910 | 6 | G/C | 32604403 | None | *HLA-DRB1* | South Asian | 0.64 | 0.43 | $1.76x10^{-11}$ | 2.23 (1.77-2.82) | $1.30x10^{-4}$ | 1.68 (1.29-2.21) | NA | |
| rs114032596 | 10 | C/T | 28810849 | non-coding intronic | *LINC01517* | South Asian | 0.06 | 0.00 | $1.83x10^{-8}$ | 9.81 (4.43-21.74) | NA | | NA | |

† Conditional analysis performed with rs17843604.
†† Conditional analysis performed with rs17843604 and rs3129713.
*** Significant SNP after Bonferroni p-value correction.

#### Supplementary Table S3: Genome wide suggestive loci associated with SSNS

| SNP | Chrom | Allele | Position | Type | Gene/ Closest gene | Allele frequency: cases A2 | Allele Frequency: controls A2 | Allelic P-value | OR (95% CI) | Replicati: Bristol vs UKBB P-value | Replicati: Bristol vs UKBB OR (95% CI) | Replicati: CureGN vs gnomAD P-value | Replicati: CureGN vs gnomAD OR (95% CI) |
|---|---|---|---|---|---|---|---|---|---|---|---|---|---|
| rs62072970 | 17 | G/A | 56125448 | Protein-coding | ANKFN1 | 0.02 | 0.01 | $1.15x10^{-7}$ | 5.08 (2.77-9.27) | 0.96 | 1.09 (0.61-1.79) | NA | |
| rs140718574 | 8 | A/T | 109898199 | Protein-coding | KCNV1 | 0.03 | 0.01 | $1.54x10^{-7}$ | 3.68 (2.26-5.98) | NA | | NA | |
| rs10098704 | 8 | A/G | 107771874 | Protein-coding | RSPO2 | 0.08 | 0.15 | $1.95x10^{-7}$ | 0.55 (0.44-0.69) | 0.04 | 0.81 (0.61-1.06) | 0.17 | 1.26 (0.91-1.70) |
| rs987174954 | 8 | A/T | 135891550 | non-coding intronic | LINC02055 | 0.01 | 0.00 | $4.14x10^{-7}$ | 10.64 (4.26-26.58) | NA | | NA | |
| rs12535555 | 7 | G/T | 3014907 | Protein-coding | CARD11 | 0.14 | 0.10 | $4.22x10^{-7}$ | 1.65 (1.36-2.01) | 0.49 | 1.07 (0.87-1.30) | 0.51 | 0.92 (0.68-1.22) |
| rs549980633 | 15 | A/G | 101859143 | Pseudogene | OR4F13P | 0.01 | 0.00 | $5.26x10^{-7}$ | 8.49 (3.68-19.57) | NA | | NA | |
| rs72614026 | 8 | T/C | 142322618 | Protein-coding | TSNARE1 | 0.03 | 0.09 | $8.06x10^{-7}$ | 0.42 (0.30-0.59) | 0.54 | 0.98 (0.68-1.37) | 0.85 | 0.90 (0.51-1.46) |
| rs1478763389 | 10 | A/G | 45904808 | Non-coding intronic | PARGP1 | 0.01 | 0.00 | $9.41x10^{-7}$ | 11.44 (4.32-30.31) | NA | | NA | |
| rs185225301 | 1 | T/C | 218141640 | snRNA | RNU1-141P | 0.01 | 0.00 | $9.84x10^{-7}$ | 6.12 (2.96-12.65) | NA | | NA | |

#### Supplementary Table S4: Genome wide suggestive loci associated with SRNS

| SNP | Chrom | Allele A1/A2 | Position | Type | Gene/ Closest gene | Allele frequency: cases A2 | Allele Frequency: controls A2 | Allelic P-value | OR (95% CI) | Replication: CureGN vs gnomAD P-value | Replication: CureGN vs gnomAD OR (95% CI) |
|---|---|---|---|---|---|---|---|---|---|---|---|
| rs58384577 | 22 | T/C | 36267167 | Protein coding (Intronic) | *APOL1* | 0.18 | 0.08 | $4.12x10^{-7}$ | 2.45 (1.73-3.46) | $4.98x10^{-6}$ | 3.75 (2.24-6.28) |
| rs9622363 | 22 | A/G | 36260509 | Protein coding (Intronic) | *APOL1* | 0.30 | 0.19 | $5.02x10^{-7}$ | 2.63 (1.80-3.83) | $3.52x10^{-5}$ | 3.51 (1.95-6.73) |
| rs9622362 | 22 | A/C | 36260398 | Protein coding (Intronic) | *APOL1* | 0.30 | 0.19 | $6.14x10^{-7}$ | 2.59 (1.78-3.77) | $2.93x10^{-5}$ | 3.35 (1.89-6.23) |
| rs60295735 | 22 | G/A | 36271108 | Protein coding | *APOL1* | 0.18 | 0.08 | $1.27x10^{-6}$ | 2.35 (1.66-3.33) | $1.88x10^{-5}$ | 3.47 (2.07-5.81) |
| rs73885319 | 22 | A/G | 36265860 | Protein coding G1 | *APOL1* | 0.17 | 0.08 | $2.12x10^{-7}$ | 2.32 (1.64-3.30) | $1.57x10^{-5}$ | 3.99 (2.38-6.70) |
| rs60910145 | 22 | T/G | 36265988 | Protein coding G1 | *APOL1* | 0.17 | 0.08 | $2.83x10^{-7}$ | 2.30 (1.62-3.27) | $1.83x10^{-5}$ | 3.95 (2.36-6.65) |

#### Supplementary Table S5: Association of APOL1 kidney disease high risk genotype with SSNS and SRNS

| Phenotype | High risk APOL1 genotype (G1/G1, G2/G2, G1/G2) frequency | OR (95% CI) | P-value* |
|---|---|---|---|
| SSNS | 35/148 (23.6%) | 1.04 (0.66-1.65) | 0.862 |
| SRNS | 15/41 (36.6%) | 2.62 (1.31-5.25) | 0.007 |
| Control population | 162/962 (16.8%) | - | - |

*ORs and p-values calculated with the logistic mixed model SAIGE using the same covariates as the Discovery GWAS.

#### Supplementary Table S6: 4 digits HLA alleles associations with SSNS by ancestry

A. 4-digit HLA alleles associations with SSNS in African ancestry

| CHR | SNP | BP | NMISS | OR | SE | L95 | U95 | STAT | P |
|---|---|---|---|---|---|---|---|---|---|
| **6** | HLA_DRB1*07:01 | 32546569 | 1105 | 4.023 | 0.1932 | 2.755 | 5.874 | 7.204 | 5.838e-13 |
| **6** | HLA_DQA1*02:01 | 32605190 | 1105 | 2.848 | 0.1807 | 1.999 | 4.059 | 5.794 | 6.886e-09 |
| **6** | HLA_DQB1*02:01 | 32627242 | 1105 | 2.406 | 0.1544 | 1.778 | 3.256 | 5.688 | 1.287e-08 |
| **6** | HLA_DQB1*06:02 | 32627268 | 1105 | 0.354 | 0.2074 | 0.2358 | 0.5315 | -5.008 | 5.503e-07 |
| **6** | HLA_DQA1*01:02 | 32605185 | 1105 | 0.5403 | 0.1525 | 0.4007 | 0.7286 | -4.036 | 5.427e-05 |
| **6** | HLA_DRB1*16:02 | 32546625 | 1105 | 5.146 | 0.4532 | 2.117 | 12.51 | 3.615 | 0.0003005 |
| **6** | HLA_DRB1*15:03 | 32546619 | 1105 | 0.4697 | 0.2155 | 0.3079 | 0.7167 | -3.506 | 0.0004554 |
| **6** | HLA_DRB1*08:04 | 32546575 | 1105 | 1.962 | 0.2291 | 1.252 | 3.074 | 2.941 | 0.003269 |
| **6** | HLA_DPB1*17:01 | 33043744 | 1105 | 1.822 | 0.2046 | 1.22 | 2.721 | 2.931 | 0.003375 |
| **6** | HLA_A*36:01 | 29910331 | 1105 | 0.3298 | 0.3814 | 0.1562 | 0.6965 | -2.908 | 0.003634 |
| **6** | HLA_DPB1*04:02 | 33043712 | 1105 | 0.5605 | 0.2363 | 0.3527 | 0.8907 | -2.45 | 0.0143 |
| **6** | HLA_DPB1*02:01 | 33043706 | 1105 | 0.5902 | 0.2158 | 0.3867 | 0.901 | -2.443 | 0.01455 |
| **6** | HLA_DRB1*11:01 | 32546584 | 1105 | 0.4997 | 0.2943 | 0.2807 | 0.8897 | -2.357 | 0.01841 |
| **6** | HLA_DRB1*12:01 | 32546593 | 1105 | 0.29 | 0.532 | 0.1022 | 0.8227 | -2.327 | 0.01998 |
| **6** | HLA_A*33:01 | 29910323 | 1105 | 2.799 | 0.4479 | 1.163 | 6.734 | 2.298 | 0.02158 |
| **6** | HLA_DQA1*01:01 | 32605184 | 1105 | 0.5912 | 0.2407 | 0.3689 | 0.9476 | -2.183 | 0.02901 |
| **6** | HLA_DRB1*13:02 | 32546597 | 1105 | 0.4737 | 0.3454 | 0.2407 | 0.932 | -2.164 | 0.03049 |
| **6** | HLA_DPB1*13:01 | 33043730 | 1105 | 0.27 | 0.6124 | 0.08131 | 0.8969 | -2.138 | 0.03254 |
| **6** | HLA_DPA1*02:01 | 33032355 | 1105 | 1.347 | 0.141 | 1.022 | 1.776 | 2.113 | 0.03461 |
| **6** | HLA_DRB1*13:01 | 32546596 | 1105 | 0.5044 | 0.3431 | 0.2575 | 0.9882 | -1.995 | 0.04609 |
| **6** | HLA_DRB1*04:05 | 32546560 | 1105 | 3.29 | 0.6001 | 1.015 | 10.66 | 1.984 | 0.04721 |
| **6** | HLA_DPB1*01:01 | 33043704 | 1105 | 1.292 | 0.131 | 0.9994 | 1.67 | 1.956 | 0.05052 |
| **6** | HLA_C*16:01 | 31236578 | 1105 | 1.477 | 0.2039 | 0.9901 | 2.202 | 1.911 | 0.05595 |

B. 4-digit HLA alleles associations with SSNS in South Asian ancestry

| CHR | SNP | BP | NMISS | OR | SE | L95 | U95 | STAT | P |
|---|---|---|---|---|---|---|---|---|---|
| **6** | HLA_DQB1*02:01 | 32627242 | 1064 | 2.583 | 0.1327 | 1.992 | 3.35 | 7.152 | 8.582e-13 |
| **6** | HLA_DRB1*07:01 | 32546569 | 1064 | 2.214 | 0.1376 | 1.691 | 2.9 | 5.776 | 7.664e-09 |
| **6** | HLA_DQA1*02:01 | 32605190 | 1064 | 2.006 | 0.1322 | 1.548 | 2.6 | 5.266 | 1.391e-07 |
| **6** | HLA_B*44:03 | 31321744 | 1064 | 2.345 | 0.1634 | 1.703 | 3.23 | 5.217 | 1.814e-07 |
| **6** | HLA_C*07:01 | 31236552 | 1064 | 2.145 | 0.1535 | 1.588 | 2.898 | 4.973 | 6.592e-07 |
| **6** | HLA_DRB1*10:01 | 32546582 | 1064 | 0.1971 | 0.3704 | 0.09537 | 0.4074 | -4.384 | 1.166e-05 |
| **6** | HLA_DQA1*05:01 | 32605196 | 1064 | 2.178 | 0.1808 | 1.528 | 3.104 | 4.304 | 1.677e-05 |
| **6** | HLA_DQA1*01:01 | 32605184 | 1064 | 0.461 | 0.1872 | 0.3194 | 0.6654 | -4.136 | 3.528e-05 |
| **6** | HLA_DQB1*05:01 | 32627261 | 1064 | 0.4234 | 0.2279 | 0.2709 | 0.6618 | -3.771 | 0.0001627 |
| **6** | HLA_DPB1*17:01 | 33043744 | 1064 | 3.802 | 0.3715 | 1.836 | 7.875 | 3.595 | 0.0003244 |
| **6** | HLA_DRB1*11:01 | 32546584 | 1064 | 2.798 | 0.3035 | 1.544 | 5.073 | 3.39 | 0.0006989 |
| **6** | HLA_DQA1*01:02 | 32605185 | 1064 | 0.5683 | 0.1733 | 0.4046 | 0.7982 | -3.26 | 0.001113 |
| **6** | HLA_B*07:02 | 31321650 | 1064 | 0.3399 | 0.3454 | 0.1727 | 0.6689 | -3.124 | 0.001783 |
| **6** | HLA_DRB1*12:02 | 32546594 | 1064 | 0.3194 | 0.3729 | 0.1538 | 0.6634 | -3.06 | 0.002211 |
| **6** | HLA_DQB1*06:03 | 32627269 | 1064 | 0.3394 | 0.4004 | 0.1548 | 0.7439 | -2.699 | 0.006954 |
| **6** | HLA_DPB1*03:01 | 33043709 | 1064 | 0.5242 | 0.2449 | 0.3244 | 0.8471 | -2.637 | 0.008355 |
| **6** | HLA_DQB1*04:02 | 32627259 | 1064 | 3.335 | 0.4633 | 1.345 | 8.268 | 2.6 | 0.009335 |
| **6** | HLA_A*02:01 | 29910252 | 1064 | 2.054 | 0.2773 | 1.193 | 3.537 | 2.597 | 0.009415 |
| **6** | HLA_DRB1*13:01 | 32546596 | 1064 | 0.3118 | 0.4524 | 0.1285 | 0.7567 | -2.576 | 0.009985 |
| **6** | HLA_B*15:02 | 31321665 | 1064 | 0.4215 | 0.3384 | 0.2171 | 0.8181 | -2.553 | 0.01067 |

C. 4-digit HLA alleles associations with SSNS in European ancestry

| CHR | SNP | BP | NMISS | OR | SE | L95 | U95 | STAT | P |
|---|---|---|---|---|---|---|---|---|---|
| 6 | HLA_DQB1*02:01 | 32627242 | 909 | 3.276 | 0.1605 | 2.392 | 4.487 | 7.392 | 1.441e-13 |
| 6 | HLA_DRB1*07:01 | 32546569 | 909 | 3.09 | 0.1744 | 2.195 | 4.349 | 6.468 | 9.962e-11 |
| 6 | HLA_DQA1*02:01 | 32605190 | 909 | 2.898 | 0.1684 | 2.083 | 4.032 | 6.317 | 2.662e-10 |
| 6 | HLA_DQA1*01:02 | 32605185 | 909 | 0.2073 | 0.3001 | 0.1151 | 0.3733 | -5.243 | 1.576e-07 |
| 6 | HLA_DRB1*15:01 | 32546617 | 909 | 0.1019 | 0.4691 | 0.04065 | 0.2556 | -4.868 | 1.129e-06 |
| 6 | HLA_DQB1*06:02 | 32627268 | 909 | 0.1083 | 0.468 | 0.0433 | 0.2711 | -4.749 | 2.043e-06 |
| 6 | HLA_DPA1*01:03 | 33032347 | 909 | 0.555 | 0.1553 | 0.4094 | 0.7525 | -3.792 | 0.0001495 |
| 6 | HLA_C*16:01 | 31236578 | 909 | 2.533 | 0.2723 | 1.485 | 4.319 | 3.413 | 0.0006437 |
| 6 | HLA_DPA1*02:01 | 33032355 | 909 | 1.731 | 0.1632 | 1.257 | 2.383 | 3.362 | 0.0007733 |
| 6 | HLA_DQA1*01:01 | 32605184 | 909 | 0.3508 | 0.3125 | 0.1901 | 0.6471 | -3.353 | 0.0008 |
| 6 | HLA_DQA1*05:01 | 32605196 | 909 | 1.739 | 0.1679 | 1.252 | 2.417 | 3.297 | 0.000978 |
| 6 | HLA_B*44:03 | 31321744 | 909 | 2.19 | 0.2463 | 1.352 | 3.55 | 3.183 | 0.001457 |
| 6 | HLA_B*07:02 | 31321650 | 909 | 0.4483 | 0.2623 | 0.2681 | 0.7497 | -3.058 | 0.002225 |
| 6 | HLA_DPB1*11:01 | 33043726 | 909 | 2.771 | 0.3338 | 1.44 | 5.331 | 3.053 | 0.002265 |
| 6 | HLA_C*07:02 | 31236553 | 909 | 0.4745 | 0.2459 | 0.293 | 0.7685 | -3.031 | 0.002439 |
| 6 | HLA_B*08:01 | 31321655 | 909 | 2.018 | 0.2353 | 1.272 | 3.2 | 2.982 | 0.002859 |
| 6 | HLA_DRB1*03:01 | 32546553 | 909 | 1.868 | 0.2115 | 1.234 | 2.827 | 2.953 | 0.003144 |
| 6 | HLA_A*30:01 | 29910308 | 909 | 4.77 | 0.5336 | 1.676 | 13.57 | 2.928 | 0.003415 |
| 6 | HLA_DQB1*05:01 | 32627261 | 909 | 0.385 | 0.3261 | 0.2031 | 0.7295 | -2.927 | 0.003423 |
| 6 | HLA_DQB1*06:03 | 32627269 | 909 | 0.2463 | 0.5212 | 0.08868 | 0.6842 | -2.688 | 0.007186 |
| 6 | HLA_DQA1*01:03 | 32605186 | 909 | 0.2878 | 0.4675 | 0.1151 | 0.7193 | -2.665 | 0.007706 |
| 6 | HLA_DRB1*13:01 | 32546596 | 909 | 0.2088 | 0.5972 | 0.06478 | 0.6731 | -2.623 | 0.00872 |
| 6 | HLA_B*44:02 | 31321743 | 909 | 0.4437 | 0.3104 | 0.2415 | 0.8152 | -2.618 | 0.00884 |
| 6 | HLA_C*08:02 | 31236561 | 909 | 2.194 | 0.3091 | 1.197 | 4.022 | 2.542 | 0.01101 |

#### Supplementary Table S7: HLA haplotype association with SSNS stratified by ancestry

A: Classic HLA 3D haplotype associated with SSNS in African ancestry

| HLA Haplotype DRB1~DQA1~DQB1 | Frequency cases | Frequency controls | Odds Ratio (95% CI) | P value |
|---|---|---|---|---|
| 07:01~02:01~02:02* | 0.20 | 0.07 | 3.24 (2.3-4.6) | $8.1 \times 10^{-13}$ |
| 08:04~05:05~03:01 | 0.04 | 0.01 | 3.21 (1.5-6.5) | 0.0003 |
| 16:02~01:02~05:02 | 0.03 | 0.01 | 2.70 (1.1-6.2) | 0.0100 |
| 03:01~05:01~02:01* | 0.09 | 0.05 | 1.75 (1.1-2.8) | 0.0124 |
| 15:03~01:02~06:02 | 0.08 | 0.13 | 0.60 (0.4-0.9) | 0.0192 |
| 11:01~05:05~03:19 | 0.01 | 0.03 | 0.26 (0.03-1.0) | 0.0444 |
| 10:01~01:05~05:01 | 0.01 | 0.03 | 0.25 (0.03-1.0) | 0.0407 |
| 13:04~05:05~03:19 | 0.00 | 0.02 | 0.00 (0.0-0.6) | 0.0132 |
| 12:01~05:05~03:01 | 0.00 | 0.01 | 0.00 (0.0-1.0) | 0.0479 |

* Significant haplotype for all ancestries.

B: Classic HLA 3D haplotype associated with SSNS in South Asian ancestry

| HLA Haplotype DRB1~DQA1~DQB1 | Frequency cases | Frequency controls | Odds Ratio (95% CI) | P value |
|---|---|---|---|---|
| 07:01~02:01~02:02* | 0.29 | 0.11 | 3.13 (2.5-4.0) | $2.2 \times 10^{-16}$ |
| 04:05~03:03~04:01 | 0.02 | 0.00 | 3.23 (1.1-10.1) | 0.0122 |
| 04:01~03:01~03:02 | 0.03 | 0.01 | 1.93 (0.9-3.9) | 0.0451 |
| 03:01~05:01~02:01* | 0.09 | 0.07 | 1.45 (1.0-2.1) | 0.0285 |
| 15:01~01:03~06:01 | 0.02 | 0.04 | 0.39 (0.2-0.8) | 0.0036 |
| 15:01~01:02~06:02 | 0.01 | 0.02 | 0.38 (0.1-1.0) | 0.0397 |
| 10:01~01:05~05:01 | 0.02 | 0.05 | 0.35 (0.2-0.7) | 0.0005 |
| 12:02~06:01~03:01 | 0.01 | 0.04 | 0.32 (0.1-0.6) | 0.0007 |
| 13:01~01:03~06:03 | 0.01 | 0.06 | 0.18 (0.1-0.4) | $1.3 \times 10^{-6}$ |
| 13:02~01:02~06:04 | 0.00 | 0.01 | 0.13 (0.0-0.8) | 0.0196 |
| 10:01~01:01~05:01 | 0.00 | 0.01 | 0.00 (0.0-1.1) | 0.0446 |

* Significant haplotype for all ancestries.

C: Classic HLA 3D haplotype associated with SSNS in European ancestry

| **HLA Haplotype DRB1~DQA1~DQB1** | **Frequency cases** | **Frequency controls** | **Odds Ratio (95% CI)** | **P value** |
|---|---|---|---|---|
| 07:01~02:01~02:02* | 0.26 | 0.09 | 3.32 (2.4-4.6) | $1.9 \times 10^{-14}$ |
| 04:01~03:01~03:02 | 0.09 | 0.04 | 2.51 (1.5-4.2) | 0.0001 |
| 03:01~05:01~02:01* | 0.22 | 0.11 | 2.17 (1.5-3.0) | $2.5 \times 10^{-6}$ |
| 01:01~01:01~05:01 | 0.04 | 0.11 | 0.32 (0.2-0.6) | 0.0003 |
| 13:01~01:03~06:03 | 0.01 | 0.06 | 0.18 (0.0-0.6) | 0.0012 |
| 14:54~01:04~05:03 | 0.00 | 0.02 | 0.17 (0.0-1.0) | 0.0473 |
| 15:01~01:02~06:02 | 0.02 | 0.12 | 0.13 (0.0-0.3) | $2.5 \times 10^{-7}$ |

* Significant haplotype for all ancestries.

#### Supplementary Table S8: Extended 6 loci HLA haplotype associated with SSNS in a large multiethnic cohort

| **HLA Haplotype A~B~C~DRB1~DQA1~DQB1~DRB1** | **Frequency cases** | **Frequency controls** | **Odds Ratio (95% CI)** | **P value** |
|---|---|---|---|---|
| 33:03~44:03~07:06~**07:01~02:01~02:02** | 0.06 | 0.01 | 8.1 (5.7-11.5) | $2.2x10^{-16}$ |
| 24:02~44:03~07:06~**07:01~02:01~02:02** | 0.01 | 0.00 | 5.4 (2.1-14.3) | $1.5x10^{-5}$ |
| 30:01~13:02~06:02~**07:01~02:01~02:02** | 0.01 | 0.00 | 3.1 (1.7-5.4) | $2.5x10^{-5}$ |
| 33:03~57:01~06:02~**07:01~02:01~03:03** | 0.01 | 0.00 | 3.0 (1.1-7.9) | 0.001 |
| 33:03~58:01~03:02~**03:01~05:01~02:01** | 0.02 | 0.01 | 2.5 (1.6-4.0) | $3.1x10^{-5}$ |
| 29:02~44:03~16:01~**07:01~02:01~02:02** | 0.02 | 0.01 | 2.1 (1.2-3.4) | 0.004 |
| 01:01~08:01~07:01~**03:01~05:01~02:01** | 0.03 | 0.02 | 1.9 (1.3-2.7) | 0.0002 |
| 03:01~07:02~07:02~15:01~01:02~06:02 | 0.00 | 0.01 | 0.3 (0.1-0.8) | 0.011 |
| 02:01~07:02~07:02~15:01~01:02~06:02 | 0.00 | 0.01 | 0.0 (0-0.38) | 0.001 |
| 02:07~46:01~01:02~09:01~03:02~03:03 | 0.00 | 0.01 | 0.0 (0-0.38) | 0.001 |

**Supplementary Table S9: Leading amino acid residue associations with SSNS stratified by ancestry**

A. Amino acid residue associations with SSNS in African ancestry

| SNP | BP | NMISS | OR | STAT | P |
|---|---|---|---|---|---|
| AA_DRB1_13_32552123_exon2_HY | 32553395 | 1105 | 3.878 | 7.353 | 1.939e-13 |
| AA_DRB1_13_32552123_exon2_HLY | 32553389 | 1105 | 3.878 | 7.353 | 1.939e-13 |
| AA_DRB1_74_32551940_exon2_LQ | 32552664 | 1105 | 3.102 | 7.295 | 2.978e-13 |
| AA_DRB1_74_32551940_exon2_GLQ | 32552653 | 1105 | 3.102 | 7.295 | 2.978e-13 |
| AA_DRB1_11_32552129_exon2_G | 32553445 | 1105 | 4.122 | 7.259 | 3.894e-13 |
| AA_DRB1_13_32552123_exon2_Y | 32553411 | 1105 | 4.122 | 7.259 | 3.894e-13 |
| AA_DRB1_14_32552120_exon2_K | 32553334 | 1105 | 4.122 | 7.259 | 3.894e-13 |
| AA_DRB1_25_32552087_exon2_Q | 32553322 | 1105 | 4.122 | 7.259 | 3.894e-13 |
| AA_DRB1_25_32552087_exon2_R | 32553323 | 1105 | 0.2426 | -7.259 | 3.894e-13 |
| AA_DRB1_30_32552072_exon2_L | 32553269 | 1105 | 4.122 | 7.259 | 3.894e-13 |
| AA_DRB1_13_32552123_exon2_LY | 32553403 | 1105 | 4.122 | 7.259 | 3.894e-13 |
| AA_DRB1_30_32552072_exon2_AL | 32553195 | 1105 | 4.122 | 7.259 | 3.894e-13 |
| AA_DRB1_30_32552072_exon2_FL | 32553238 | 1105 | 4.122 | 7.259 | 3.894e-13 |
| AA_DRB1_30_32552072_exon2_AFL | 32553178 | 1105 | 4.122 | 7.259 | 3.894e-13 |
| AA_DRB1_74_32551940_exon2_AER | 32552613 | 1105 | 0.3265 | -7.215 | 5.377e-13 |
| AA_DRB1_74_32551940_exon2_LQx | 32552666 | 1105 | 3.063 | 7.215 | 5.377e-13 |
| AA_DRB1_74_32551940_exon2_Q | 32552671 | 1105 | 4.023 | 7.204 | 5.838e-13 |
| AA_DRB1_74_32551940_exon2_GQ | 32552656 | 1105 | 4.023 | 7.204 | 5.838e-13 |

B. Amino acid residue associations with SSNS in South Asian ancestry

| CHR | SNP | BP | NMISS | OR | STAT | P |
|---|---|---|---|---|---|---|
| 6 | AA_DQB1_71_32632639_exon2_AT | 32632645 | 1064 | 0.3757 | -7.604 | 2.871e-14 |
| 6 | AA_DQB1_28_32632768_exon2 | 32632768 | 1064 | 0.3844 | -7.202 | 5.952e-13 |
| 6 | AA_DQB1_46_32632714_exon2 | 32632714 | 1064 | 0.3844 | -7.202 | 5.952e-13 |
| 6 | AA_DQB1_47_32632711_exon2 | 32632711 | 1064 | 0.3844 | -7.202 | 5.952e-13 |
| 6 | AA_DQB1_52_32632696_exon2 | 32632696 | 1064 | 0.3844 | -7.202 | 5.952e-13 |
| 6 | AA_DQB1_30_32632762_exon2_S | 32632764 | 1064 | 2.601 | 7.202 | 5.952e-13 |
| 6 | AA_DQB1_37_32632741_exon2_I | 32632743 | 1064 | 2.601 | 7.202 | 5.952e-13 |
| 6 | AA_DQB1_55_32632687_exon2_L | 32632687 | 1064 | 2.601 | 7.202 | 5.952e-13 |
| 6 | AA_DQB1_71_32632639_exon2_K | 32632647 | 1064 | 2.601 | 7.202 | 5.952e-13 |
| 6 | AA_DQB1_74_32632630_exon2_A | 32632630 | 1064 | 2.601 | 7.202 | 5.952e-13 |
| 6 | AA_DQB1_55_32632687_exon2_LQ | 32632702 | 1064 | 2.601 | 7.202 | 5.952e-13 |

C. Amino acid residue associations with SSNS in European ancestry

| SNP | BP | NMISS | OR | STAT | P |
|---|---|---|---|---|---|
| AA_DQB1_57_32632681_exon2_A | 32632681 | 909 | 3.657 | 8.201 | 2.392e-16 |
| AA_DQB1_57_32632681_exon2_AS | 32632685 | 909 | 3.535 | 7.961 | 1.711e-15 |
| AA_DQA1_69_32609285_exon2_L | 32609293 | 909 | 4.558 | 7.833 | 4.775e-15 |
| AA_DQA1_-16_32605258_exon1_M | 32605259 | 909 | 4.558 | 7.833 | 4.775e-15 |
| AA_DQA1_69_32609285_exon2_AT | 32609290 | 909 | 0.2194 | -7.833 | 4.775e-15 |
| AA_DQB1_70_32632642_exon2_R | 32632659 | 909 | 5.06 | 7.666 | 1.774e-14 |
| AA_DQA1_11_32609111_exon2 | 32609111 | 909 | 5.032 | 7.641 | 2.156e-14 |
| AA_DQA1_18_32609132_exon2 | 32609132 | 909 | 5.032 | 7.641 | 2.156e-14 |
| AA_DQA1_45_32609213_exon2 | 32609213 | 909 | 5.032 | 7.641 | 2.156e-14 |
| AA_DQA1_48_32609222_exon2 | 32609222 | 909 | 0.1987 | -7.641 | 2.156e-14 |
| AA_DQA1_47_32609219_exon2_R | 32609241 | 909 | 0.1987 | -7.641 | 2.156e-14 |
| AA_DQA1_50_32609228_exon2_E | 32609228 | 909 | 0.1987 | -7.641 | 2.156e-14 |
| AA_DQA1_52_32609234_exon2_S | 32609253 | 909 | 0.1987 | -7.641 | 2.156e-14 |
| AA_DQA1_53_32609237_exon2_K | 32609237 | 909 | 0.1987 | -7.641 | 2.156e-14 |
| AA_DQA1_55_32609243_exon2_R | 32609265 | 909 | 5.032 | 7.641 | 2.156e-14 |
| AA_DQA1_56_32609246_exon2_G | 32609272 | 909 | 0.1987 | -7.641 | 2.156e-14 |
| AA_DQA1_61_32609261_exon2_F | 32609261 | 909 | 5.032 | 7.641 | 2.156e-14 |
| AA_DQA1_64_32609270_exon2_T | 32609281 | 909 | 5.032 | 7.641 | 2.156e-14 |
| AA_DQA1_66_32609276_exon2_I | 32609276 | 909 | 5.032 | 7.641 | 2.156e-14 |
| AA_DQA1_66_32609276_exon2_M | 32609286 | 909 | 0.1987 | -7.641 | 2.156e-14 |

**Supplementary Table S10: Leading significant amino acid residue associations with SSNS and conditional analyses by ancestry**

| | South Asian | European | African |
|---|---|---|---|
| Most significant marker (was amino acid position in each group) | AA_DQB1_71<br>$2.871 \times 10^{-14}$ | AA_DQBI_57<br>$2.392 \times 10^{-16}$ | AA_DRB1_13<br>$1.939 \times 10^{-1}3$ |
| Most significant marker conditional on leading marker | rs3104407<br>$2.883 \times 10^{-8}$ | AA_DRB1_86<br>$2.171 \times 10^{-8}$ | rs112333652<br>$8.863 \times 10^{-7}$ |
| Most significant amino acid position conditional on leading marker | AA_DRB1_32<br>$1.119 \times 10^{-6}$ | AA_DRB1_86<br>$2.171 \times 10^{-8}$ | AA_DQB1_125<br>$1.046 \times 10^{-4}$ |
| Most significant amino acid positions conditional on two leading amino acid positions | AA_DRB1_233<br>$1.112 \times 10^{-5}$ | AA_DQA1_75<br>$4.996 \times 10^{-5}$ | AA_A_9<br>3.131. X 10-4<br><br>*AA_DRB1_233<br>$3.289 \times 10^{-3}$ |

*Most significant amino acid position in MHC class II allele after two rounds of conditional analysis.

**Supplementary Table 11: Utility of the primary HLA risk haplotype and PRS as screening tests to distinguish SSNS (positive class) from SRNS**

| Predictor | | Sensitivity | Specificity | PPV | NPV | Accuracy | AUC |
|---|---|---|---|---|---|---|---|
| HLA haplotype only | | | | | | | |
| | Discovery | 100 | 0 | 79 | - | 79 | 0.612 |
| | Bristol | 100 | 0 | 67 | - | 67 | 0.574 |
| | CureGN | 100 | 0 | 60 | - | 60 | 0.618 |
| PRS-CT only | | | | | | | |
| | Discovery | 100 | 2 | 79 | 100 | 79 | 0.657 |
| | Bristol | 98 | 7 | 68 | 60 | 68 | 0.627 |
| | CureGN | 85 | 30 | 64 | 57 | 63 | 0.630 |
| PRS-LDPRED only | | | | | | | |
| | Discovery | 98 | 23 | 83 | 73 | 82 | 0.771 |
| | Bristol | 96 | 6 | 68 | 43 | 67 | 0.626 |
| | CureGN | 81 | 36 | 65 | 56 | 63 | 0.650 |
| HLA haplotype + PRS-CT | | | | | | | |
| | Discovery | 100 | 0 | 79 | - | 79 | 0.658 |
| | Bristol | 98 | 6 | 68 | 58 | 68 | 0.626 |
| | CureGN | 76 | 41 | 65 | 53 | 62 | 0.649 |
| **HLA haplotype + PRS-LDPRED** | | | | | | | |
| | Discovery | **96** | **27** | **83** | **65** | **82** | **0.794** |
| | Bristol | **96** | **6** | **68** | **43** | **67** | **0.627** |
| | CureGN | **77** | **40** | **66** | **54** | **62** | **0.656** |
| **HLA haplotype + PRS-LDPRED + 3PCs** | | | | | | | |
| | Discovery | **93** | **41** | **86** | **61** | **82** | **0.854** |
| | Bristol | **95** | **15** | **69** | **58** | **68** | **0.658** |
| | CureGN | **81** | **41** | **67** | **60** | **65** | **0.674** |
| HLA haplotype + PRS-LDPRED + age* | | | | | | | |
| | Discovery | 96 | 40 | 87 | 72 | 85 | 0.836 |
| | Bristol | 96 | 10 | 70 | 54 | 69 | 0.644 |
| | CureGN | 81 | 47 | 69 | 63 | 67 | 0.692 |
| **HLA haplotype + PRS-LDPRED + 3PCs + age*** | | | | | | | |
| | Discovery | **95** | **49** | **88** | **71** | **86** | **0.878** |
| | Bristol | **94** | **19** | **72** | **58** | **71** | **0.675** |
| | CureGN | **80** | **47** | **69** | **62** | **67** | **0.700** |

*All values expressed as percentages except AUC. Thresholded values correspond to Pr( predicted SSNS ) >= 0.5.*
**Age has missing data in the discovery cohort (for n=42 SSNS and n=22 SRNS) and in the Bristol cohort (n=1 SSNS and n=14 SRNS).*
*The "age" variable may not be consistently defined across cohorts.*

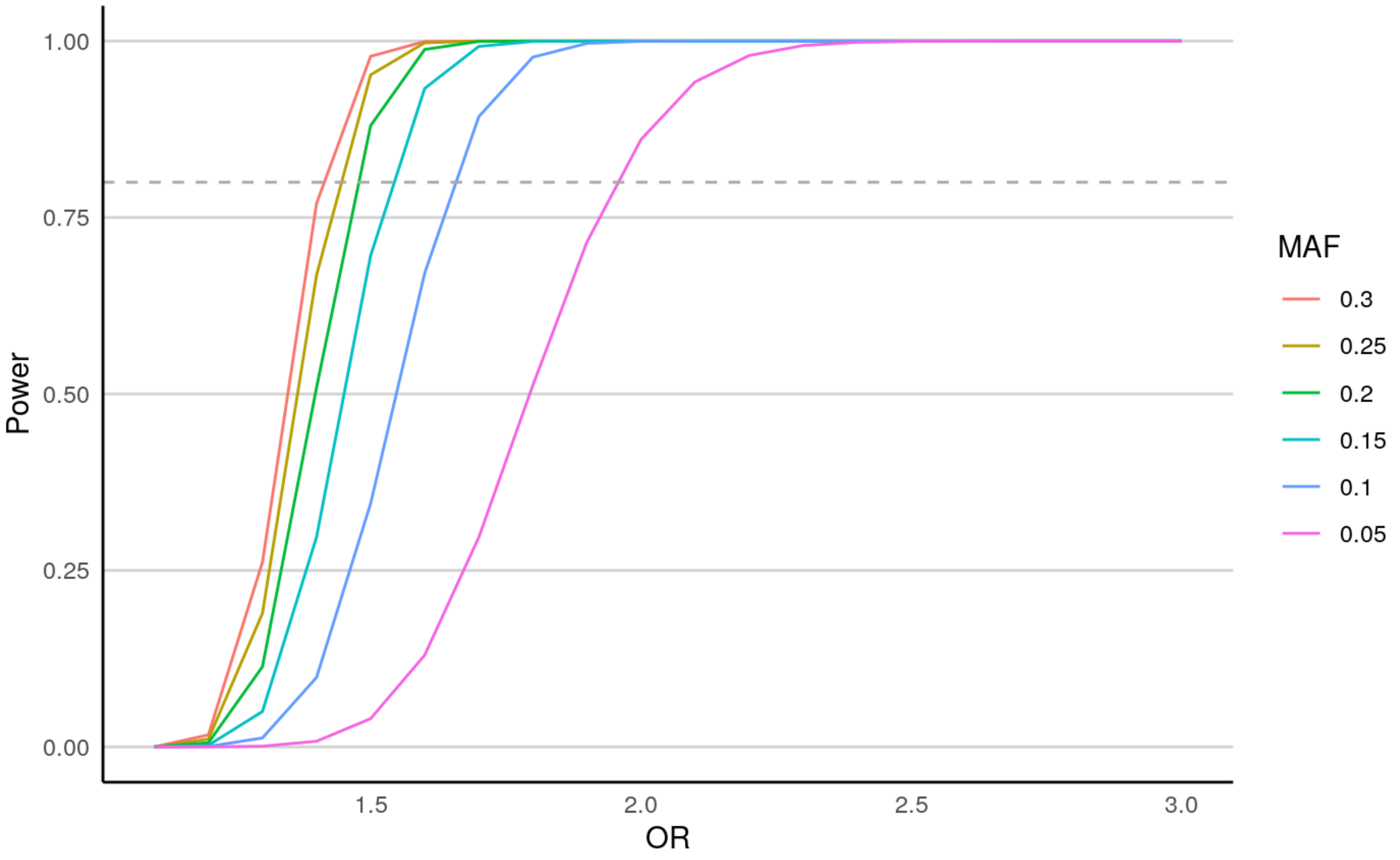


**Supplementary Figure S1. Power estimates for discovery cohort.** Power estimates for the discovery sample (994 cases and 3558 controls) for minor allele frequency (MAF) from 0.05-0.3 for alpha = $5 \times 10^{-8}$ (genome-wide significant p-value) under an additive genetic model.

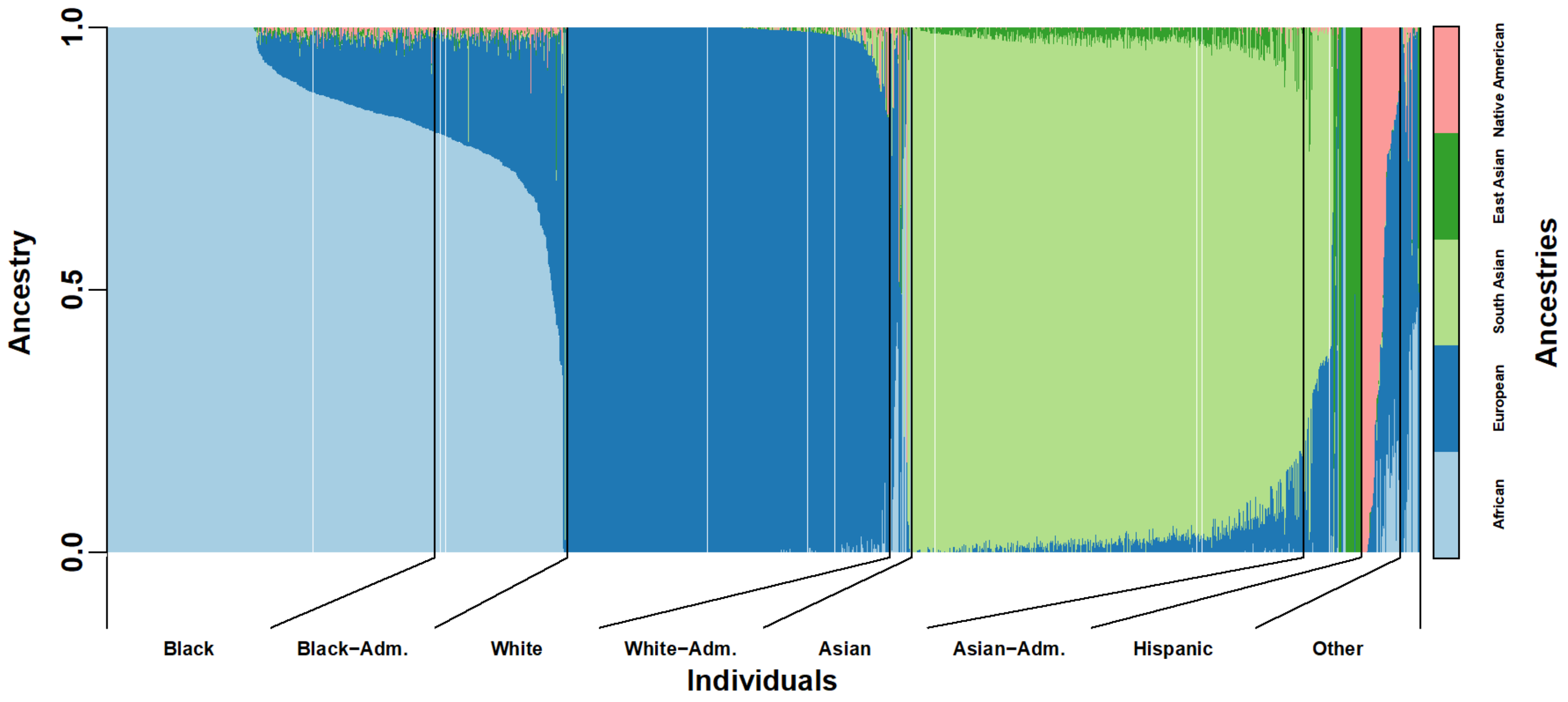


**Supplementary Figure S2. Admixture inference of GWAS discovery (genotyped by array) samples.** Individuals are grouped visually on the x-axis according to their self-described race and ethnicity, where “Other” (originally 2 individuals) also contains “Mixed” (20) and “Unknown” (8) categories. K = 5 ancestries were inferred and labeled according to their correlation to race/ethnicity labels. Black, White, and Asian individuals were then split further into those with more than 80% of the primary ancestry of the group (African, European, and South Asian, respectively), and the rest, which are highly admixed or fit other ancestry groups better, and were relabeled as Black-Adm, White-Adm, and Asian-Adm, respectively. Of note, East Asians were originally labeled as “Asian” together with the much more numerous South Asians, are separated into “Asian-Adm” along with highly admixed South Asians.

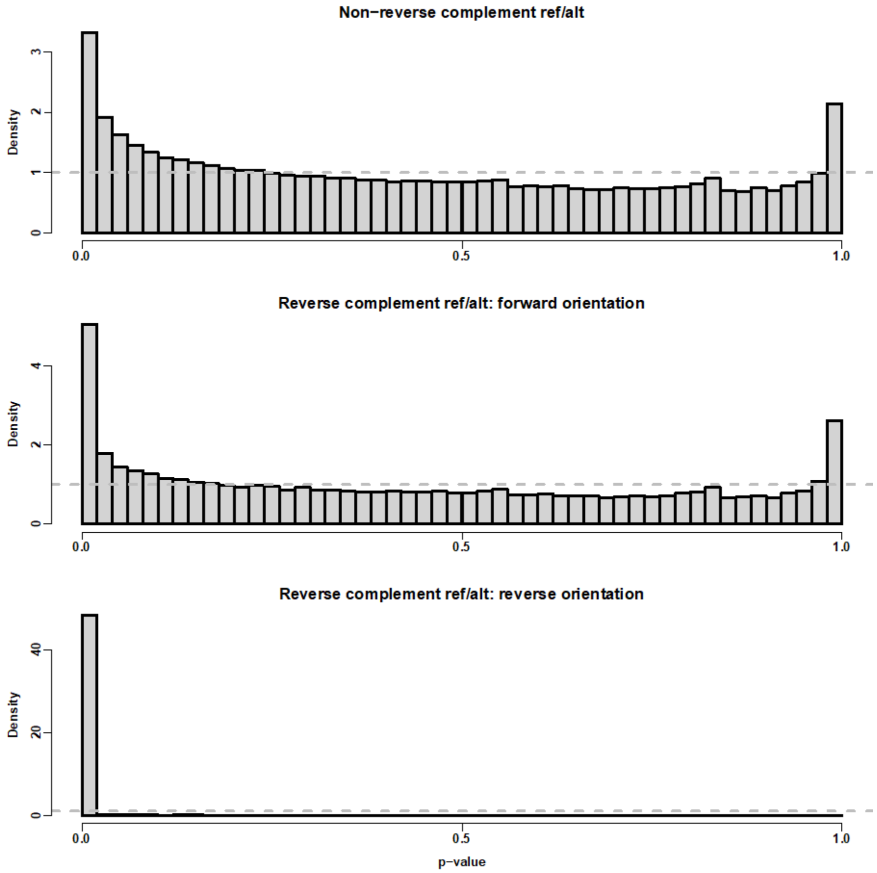


**Supplementary Figure S3. P-value distributions for platform-specific genotyping bias test.** Each p-value tests one SNP under a joint null hypothesis that there are no platform-differential genotyping biases in any of the ancestries. The null hypothesis (that there is indeed no bias) holds for most SNPs (top and middle rows), whose p-value distribution is approximately uniform. The horizontal gray dashed line marks the uniform density height, which is expected when the null hypothesis holds for all SNPs. Significant loci in this test are removed. SNPs whose reference (ref) and alternative (alt) alleles are reverse complements of each other can additionally be flipped if they were in the wrong strand. Most reverse complement cases were already in the correct orientation (middle row), as the null hypothesis was less likely to hold in the reverse orientation (more p-values are significant; bottom row). Nevertheless, 3178 SNPs were flipped, whose reverse orientation p-values were not significant and larger than their forward orientation counterparts. SNPs significant both ways are removed.

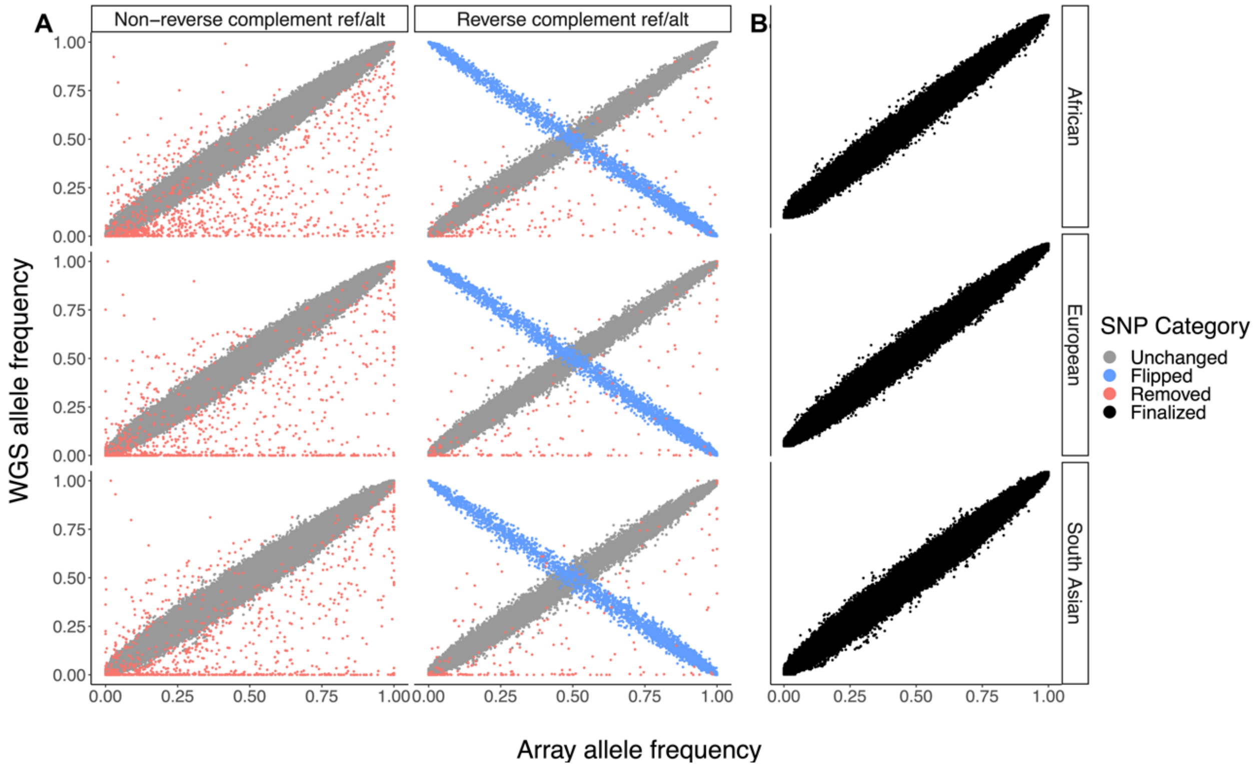


**Supplementary Figure S4. Comparison of allele frequencies and platform-differential test results.** Each point is a SNP, with control allele frequencies in Array (study-genotyped controls) and WGS (1000 Genomes) genotyping platforms along the x and y axes, respectively. (A) Each SNP is classified as Unchanged, Flipped, or Removed, depending on the outcome of the platform-specific allele frequency test (see Methods). Allele frequencies vary per ancestry (rows) but the test considers all of these ancestries jointly and results in the same classification for each SNP across ancestries. The testing procedure is more complex for SNPs whose reference (ref) and alternative (alt) alleles are reverse complements than for those that are not (columns). For non-reverse complement ref/alt SNPs (678,920), the test classifies as Unchanged (677,877) those that are consistently near the diagonal in all ancestries, and as Removed (1,043) those that are significantly far from the diagonal in at least one ancestry. For reverse complement ref/alt SNPs (83,708), most are Unchanged (80,310), but we also test if counting the opposite allele results in a better fit to the data, classified as Flipped (3,178) if they are consistently near the negative diagonal (y=1-x) in all ancestries, or Removed (220) when significantly far from both diagonals in at least one ancestry. (B) After allele frequency quality control processing based on SNP classification, we see a clear agreement in allele frequency between WGS and array controls.

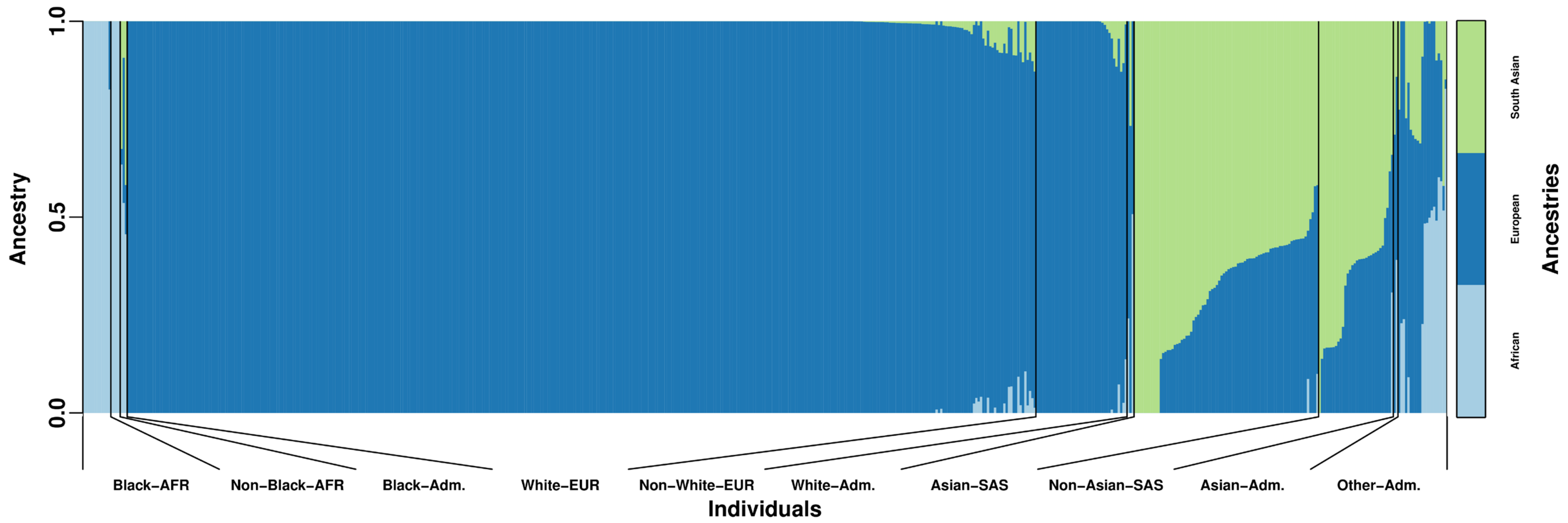


**Supplementary Figure S5. Admixture analysis of Bristol cohort.** All Bristol patients were genotyped on the same array used for the GWAS discovery dataset. This admixture analysis was done with methods similar to **Supplementary Figure S2**. In this case K = 3 produced the most interpretable results, distinguishing the most prevalent ancestries in this cohort.

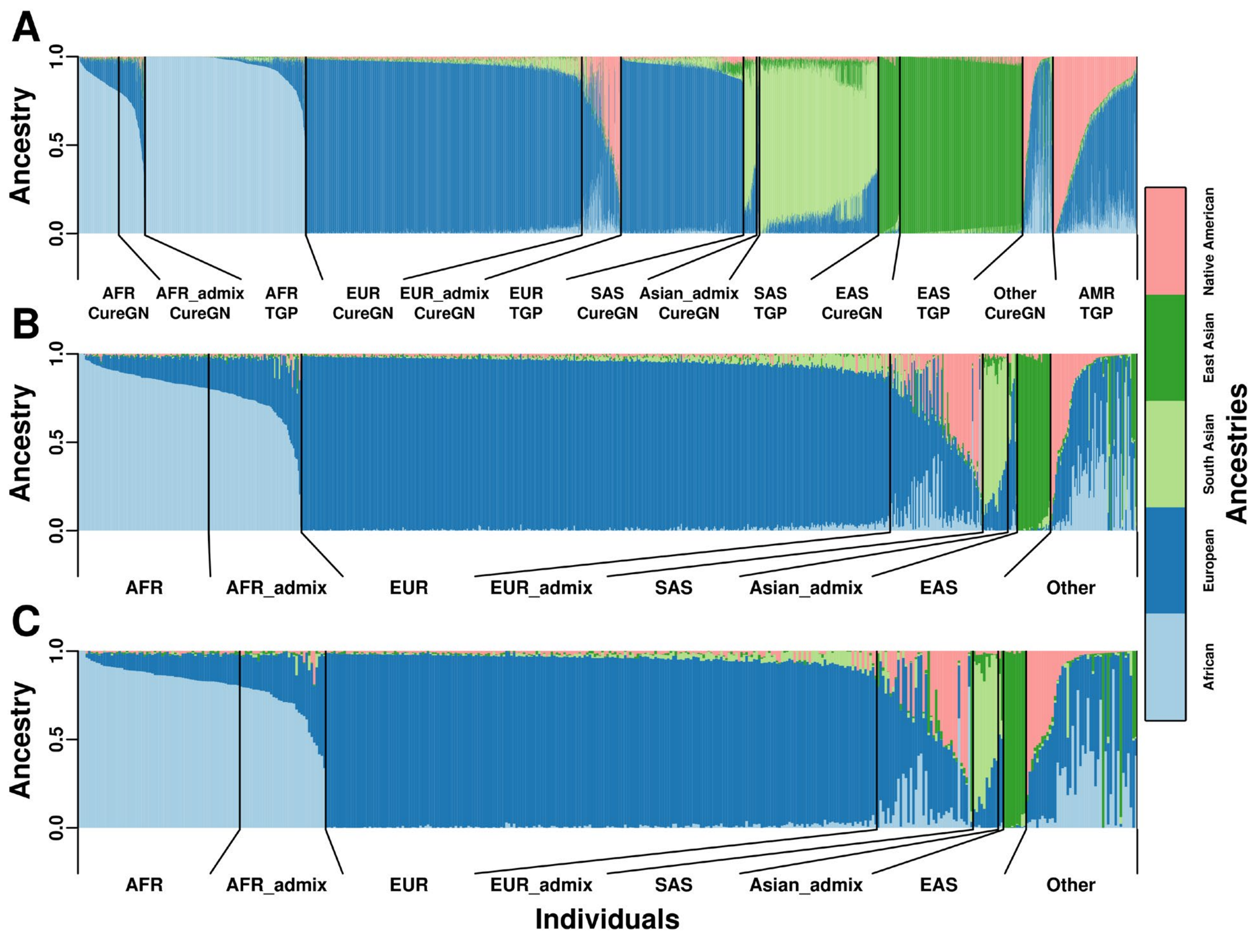


**Supplementary Figure S6. Admixture analysis of the CureGN cohort. A.** The admixture analysis was performed with the complete CureGN cohort of n=1,850 individuals (of all ages, including MCD, FSGS, as well as Membranous Nephropathy (MN) and IgA Nephropathy (IgAN) patients that were otherwise not used in our study) merged with all of the 1000 Genomes Project (TGP; n=2,504). Merging these two datasets was justified because both are whole-genome sequencing and this analysis demonstrates that no platform-specific bias is present since matching populations from both datasets had coherent admixture proportion distributions. **B.** Admixture results subset to patients treated as having Nephrotic Syndrome (MCD and FSGS patients of all ages, total n=891). **C.** Admixture results subset to patients treated as having SSNS (MCD and age of onset <=21) or SRNS (FSGS and age of onset <=21), total n= 419.

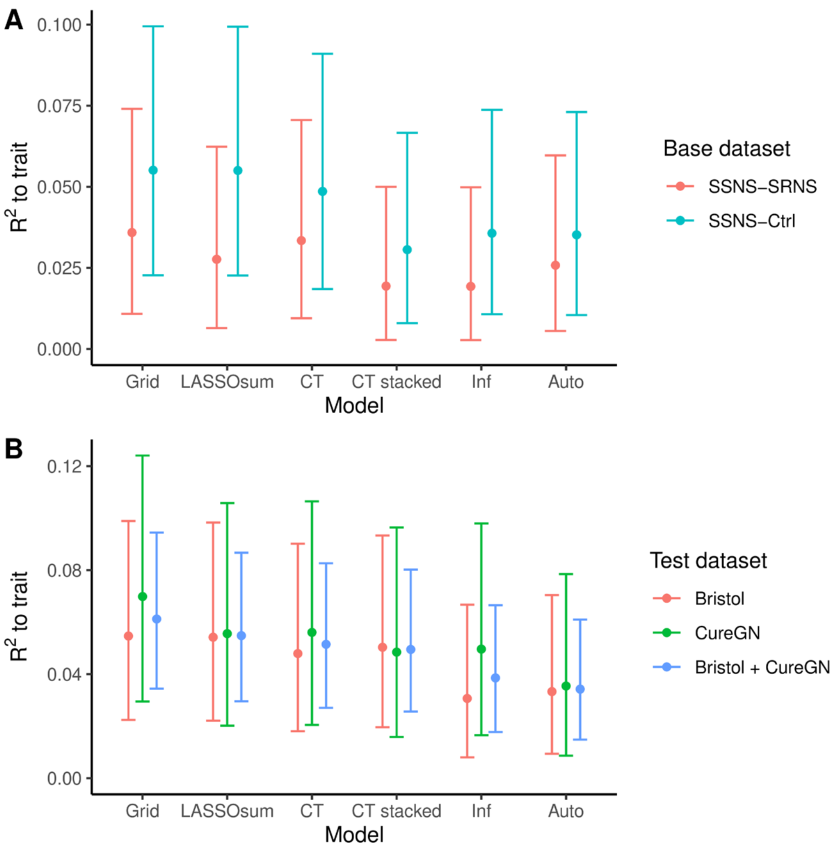


**Supplementary Figure S7. Evaluation of base data and models in testing datasets for predicting steroid therapy response.** In both panels, the 6 PRS training models on the x-axis are as implemented in the LDPred2 R package (see supplemental methods for descriptions), and are ordered roughly by performance. The y-axis shows the squared partial correlation $R^2$ to the true trait values in the test data, conditioned on the top 10 PCs (mean and 95% CIs calculated using LDPred2). **A.** PRS derived from our SSNS-vs-Control base data always perform better than SSNS-vs-SRNS base data (both from the discovery GWASes presented earlier), for every PRS training model considered. Here CureGN (SSNS-vs-SRNS trait) was used to train all PRS, and Bristol (also SSNS-vs-SRNS) was used to test the resulting PRS. Although SSNS-vs-SRNS is hypothetically better base data since the goal is to predict SSNS-vs-SRNS, in our study it has a much smaller sample size than SSNS-vs-Control base data, hence its observed advantage. **B.** Performance of PRS based on SSNS-vs-Control base data and SSNS-vs-SRNS training data, both disjoint subset of the discovery dataset, as tested on Bristol, CureGN, and the combination. We observe consistent performance in both datasets.

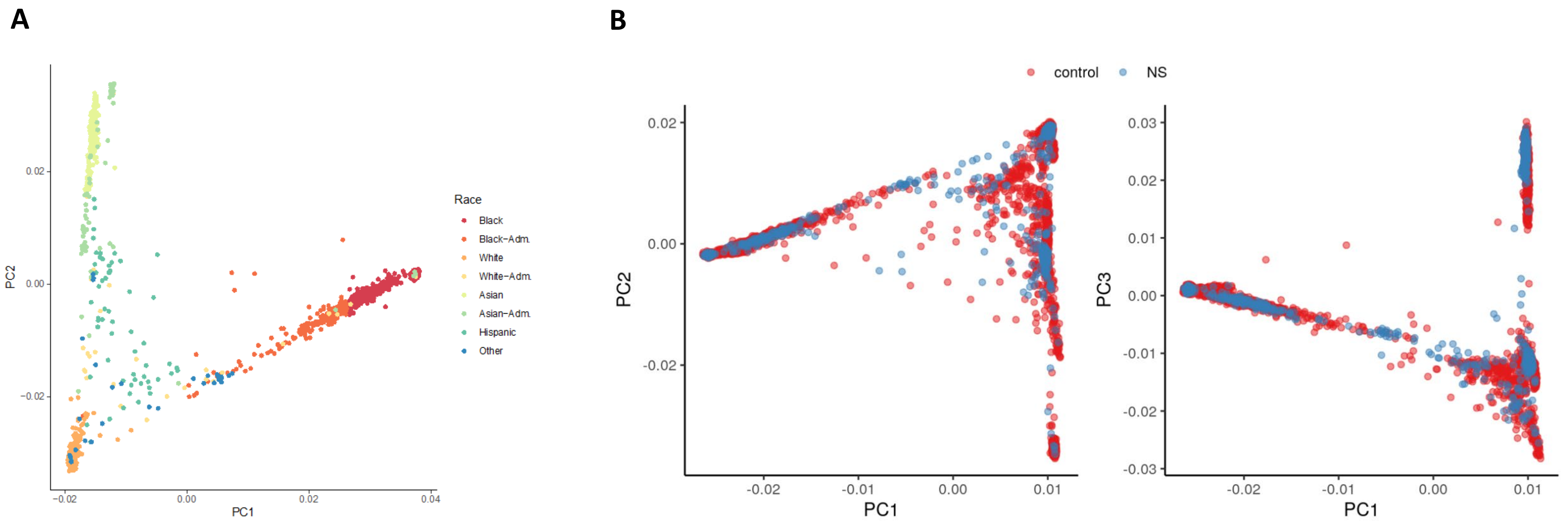


**Supplementary Figure S8. A. Admixture-informed race and ethnicity clustering on principal components space.** The unadmixed ancestry groups Black, White, and Asian form clear clusters at the vertices of the space, whereas the admixed subgroups Black-Adm, White-Adm, and Asian-Adm join Hispanic and Other in spanning the space between the three vertices. The only exception is a subcluster of Asian-Adm, which we associate with East Asian ancestry, which forms a fourth vertex adjacent to the South Asian cluster. **B. PCA of cases and controls after data harmonization and imputation.**

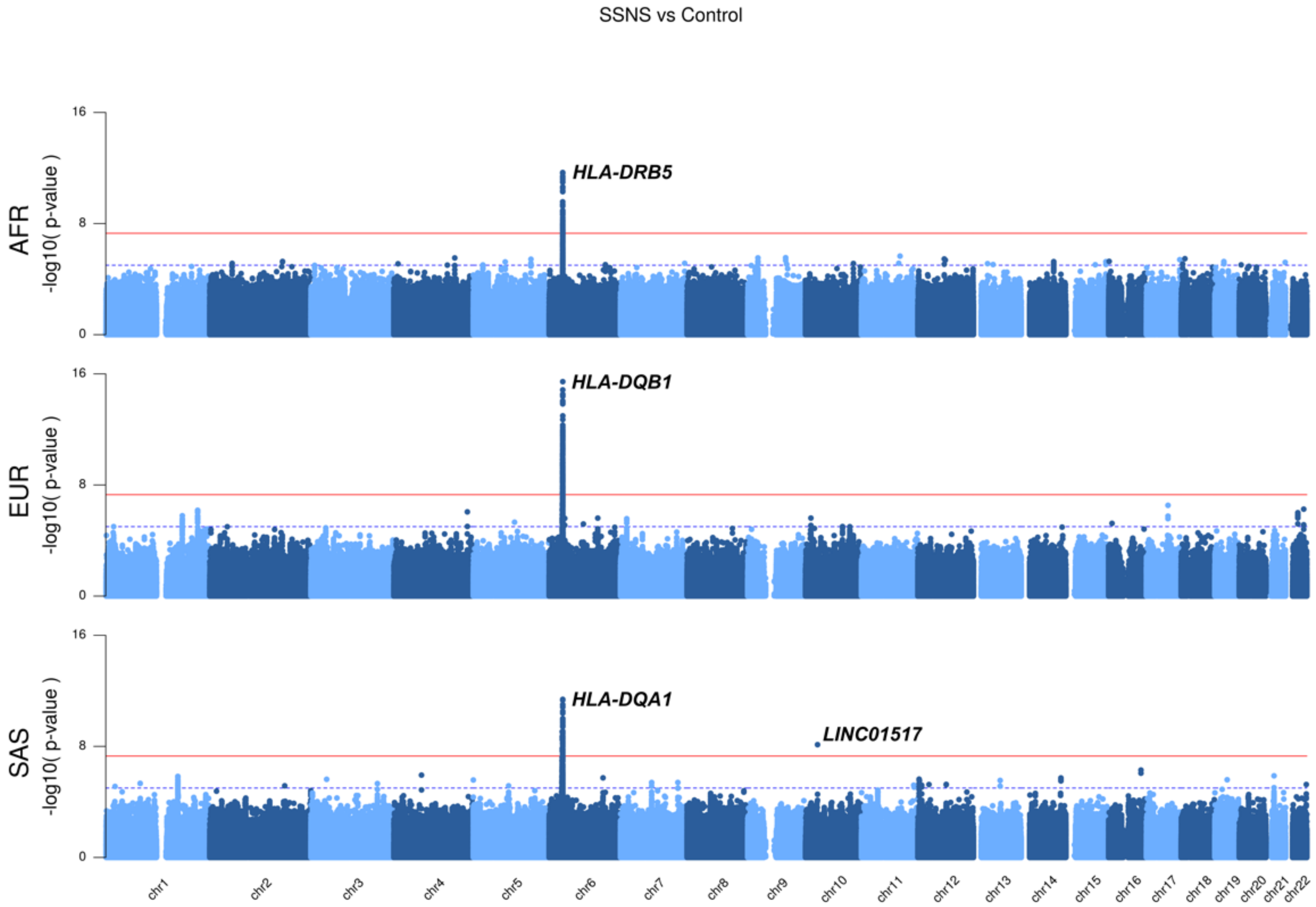


**Supplementary Figure S9: Manhattan plots of GWAS for SSNS by ancestry group.** AFR: African ancestry; EUR: European ancestry; SAS: South Asian ancestry. These ancestry groups included only individuals with 80% or more of each primary ancestry as shown in **Supplementary Figure S2**.

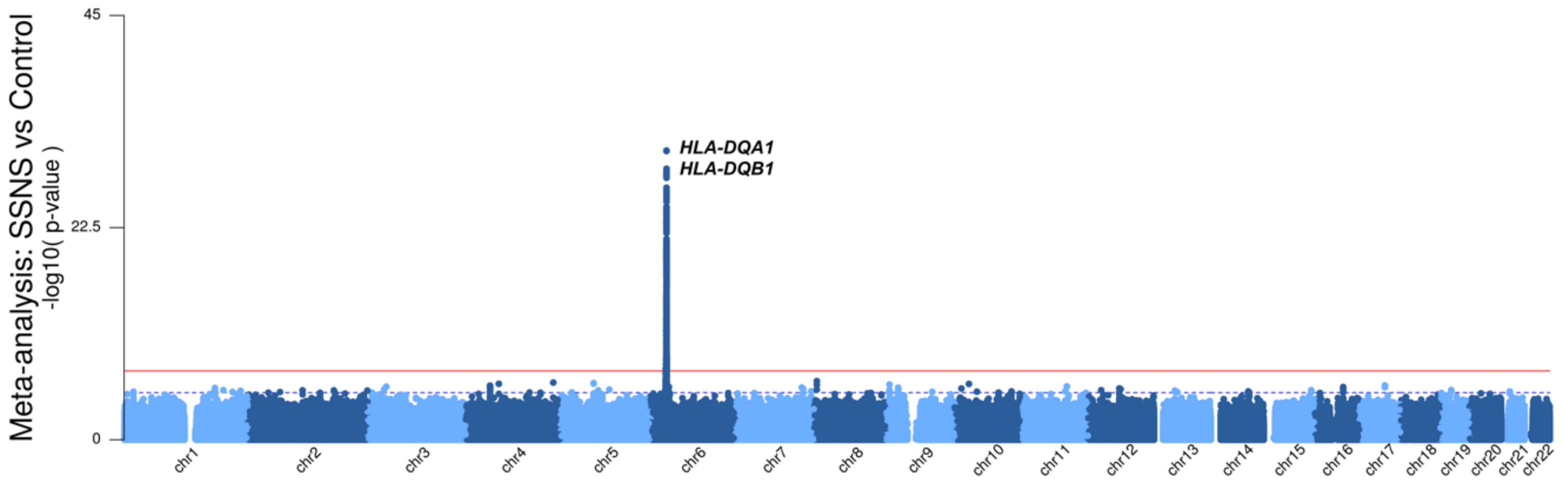


**Supplementary Figure S10: Trans-ancestry meta-analysis for SSNS.** The three ancestry-specific GWAS shown in **Supplementary Fig. S9** were meta-analyzed in this figure. Admixed individuals were excluded from all ancestry-specific GWASes as well as from this meta-analysis.

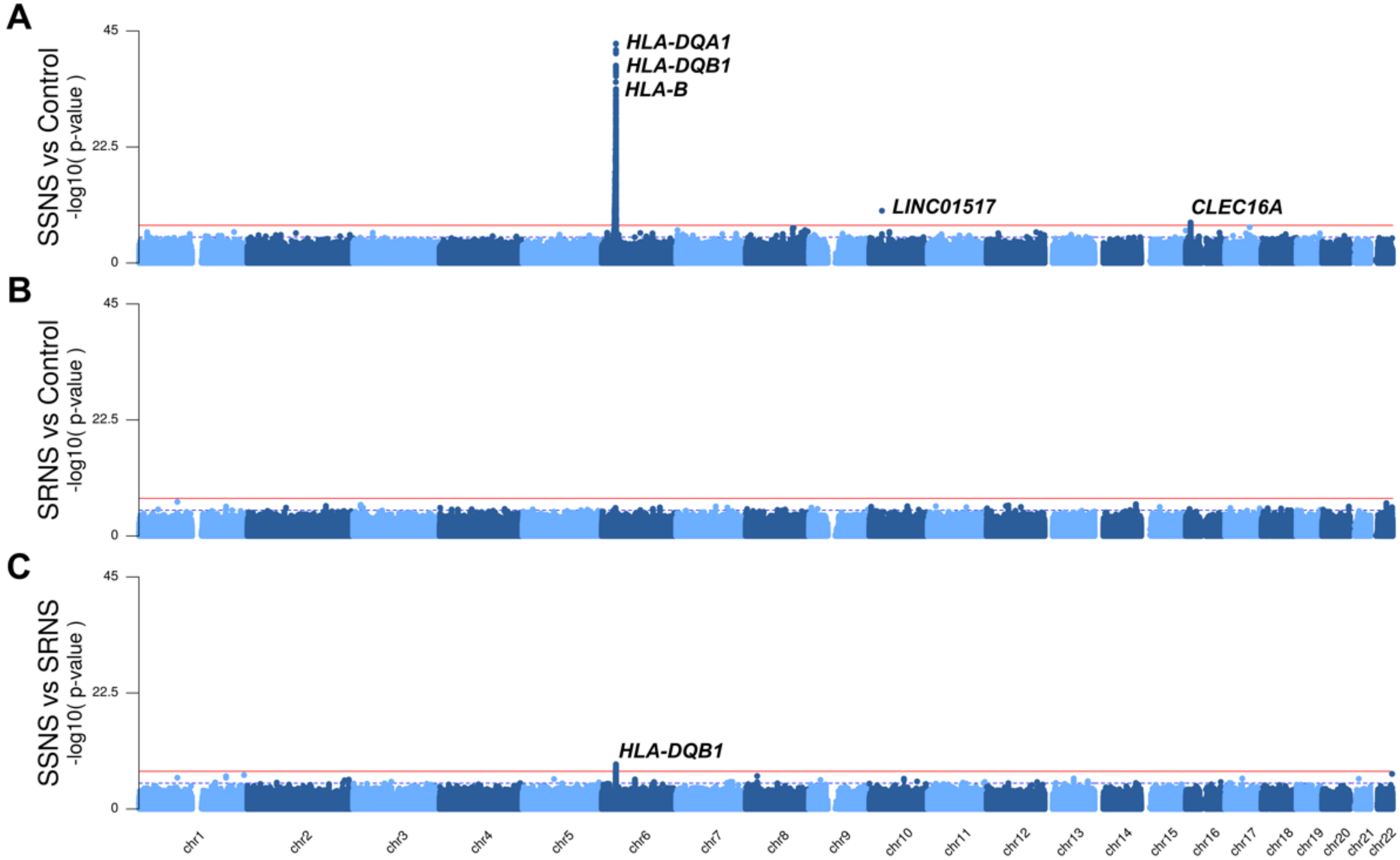


**Supplementary Figure S11: Manhattan plots of GWAS for (A) SSNS versus controls (B) SRNS versus controls, (C) SRNS versus SSNS**

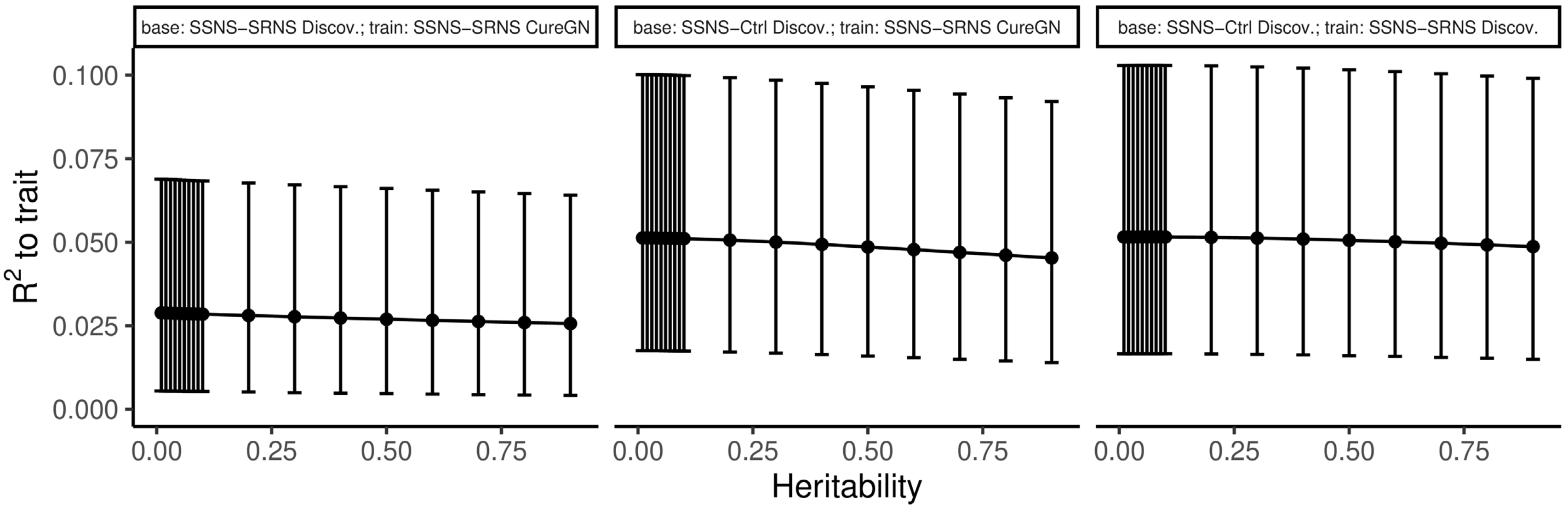


**Supplementary Figure S12. Parameter training for LDPred2 inf model.** Models are relatively insensitive to the value of the heritability ($h^2$) parameter (x-axis), but lower heritability yields slightly higher $R^2$ (y-axis). Training performance was worse (lower $R^2$ values) when the base data was SSNS-vs-SRNS compared to SSNS-vs-Control (panel columns).

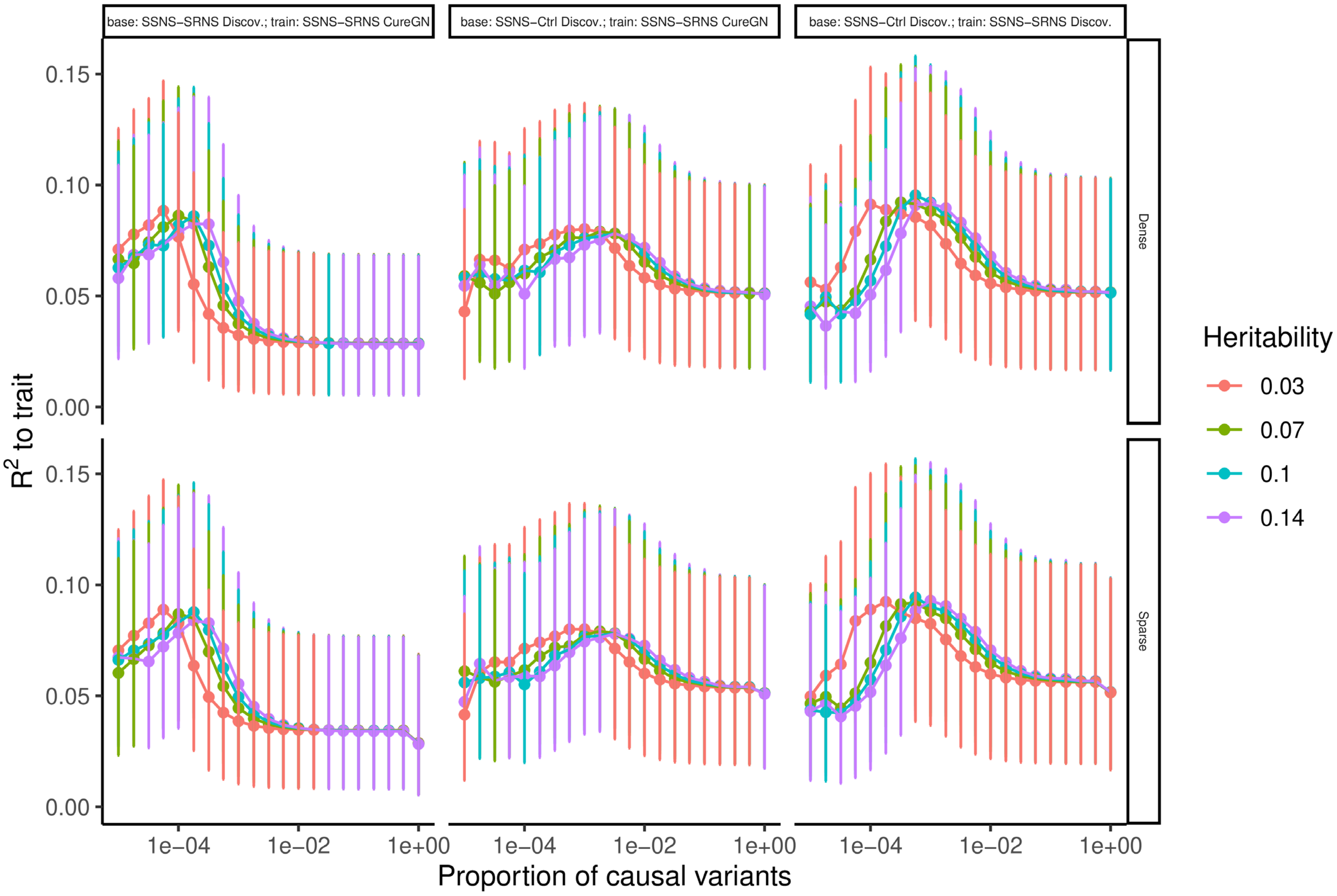


**Supplementary Figure S13. Parameter training for LDPred2 grid model.** Models are relatively insensitive to the value of the heritability ($h^2$) parameter (colors) and whether the model is sparse or dense (panel rows), but lower proportions of causal variants (x-axis) yield considerably higher performance (higher $R^2$, y-axis). Training performance was comparable when the base data was SSNS-vs-SRNS compared to SSNS-vs-Control (panel columns).

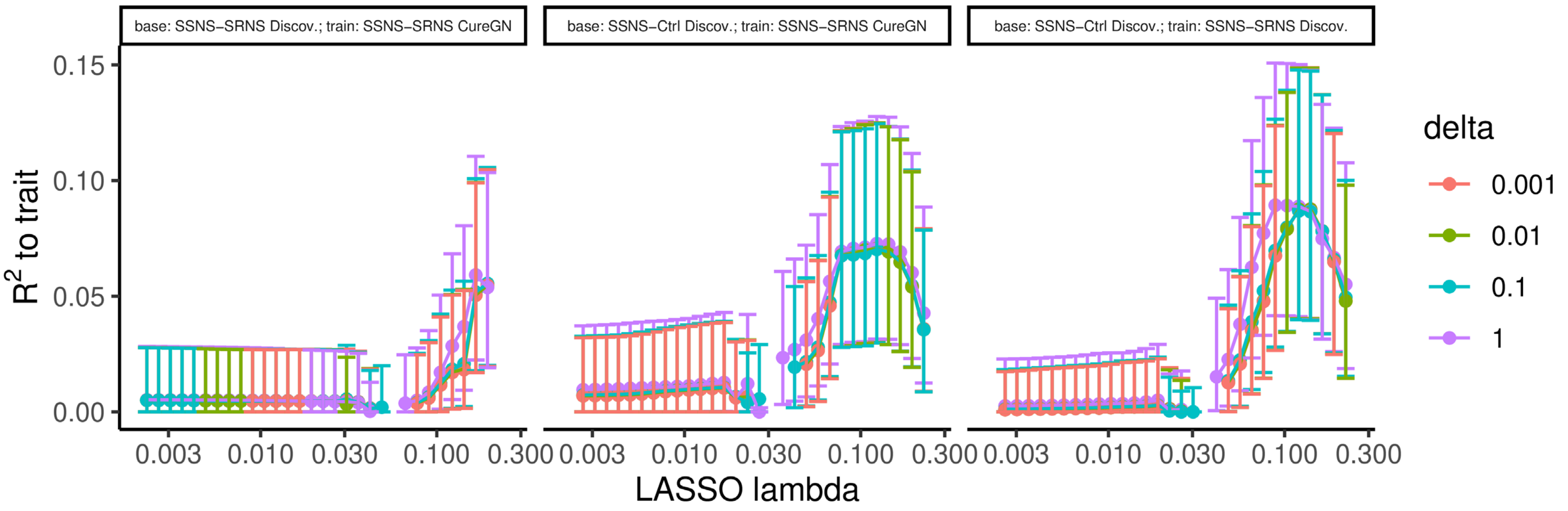


**Supplementary Figure S14. Parameter training for LASSOsum2 model.** Models are relatively insensitive to the value of the delta parameter (shrinkage; colors), but higher penalization coefficient lambda (x-axis) yielded considerably higher performance ($R^2$; y-axis). Training performance was worse when the base data was SSNS-vs-SRNS compared to SSNS-vs-Control (panel columns). Negative R values that appeared here were set to zero before squaring.

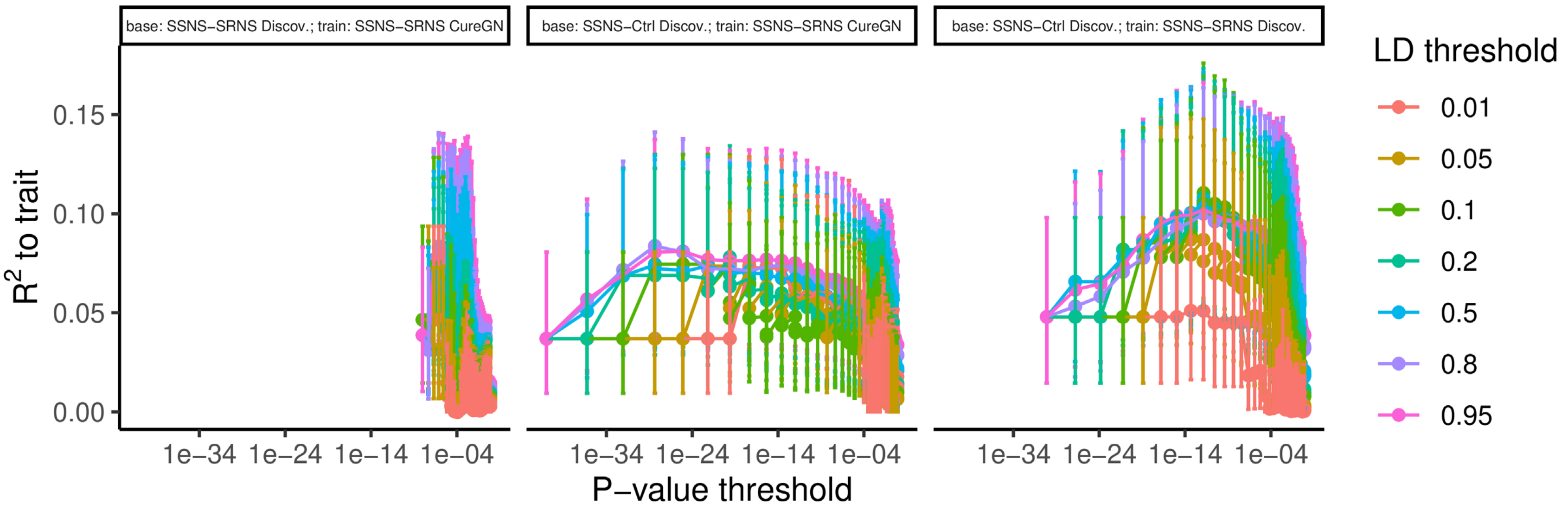


**Supplementary Figure S15. Parameter training for clump and threshold (C+T) model.** Model performance ($R^2$, y-axis) is sensitive to both the LD (colors) and p-value thresholds (x-axis). When the base data is SSNS-vs-SRNS (first panel) very small p-values are not achieved, explaining why the curve is concentrated to the right compared to the other two panels. Negative R values that appeared here were set to zero before squaring.

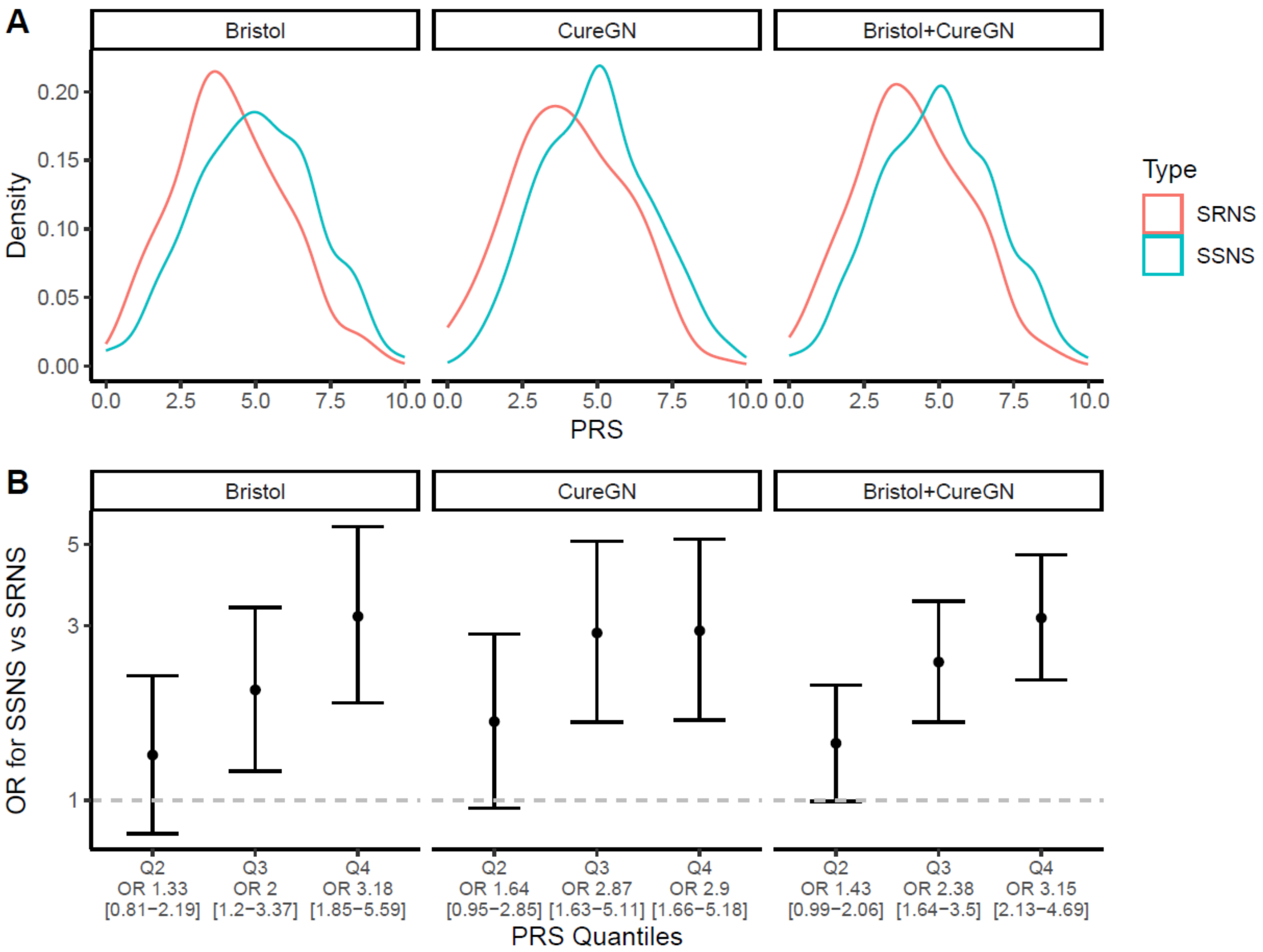


**Supplementary Figure S16. Clump and Threshold (C+T) performs similarly to LDPred2 grid method on testing datasets.** Same setup as **Figure 3**, which shows the results for LDPred2 grid method, but here it was repeated for PRS developed using the C+T method. One big difference is the scale of the PRS is different, going between roughly 0 and 10 for C+T, whereas they ranged roughly between 0 and 2 for LDPred2. There is also more overlap between the ORs and CIs of for the top two PRS quartiles in CureGN and in the combined dataset, compared to the LDPred2 results.
